# A contextual genomic perspective on physical activity and its relationship to health, well-being and illness

**DOI:** 10.1101/2025.04.10.25324713

**Authors:** Marco Galimberti, Daniel F. Levey, Joseph D. Deak, Keyrun Adhikari, Cassie Overstreet, Priya Gupta, Rachana Nitin, Hang Zhou, Nicole J. Lake, Kelly M. Harrington, Luc Djousse, Lea K. Davis, VA Million Veteran Program, J. Michael Gaziano, Murray B. Stein, Joel Gelernter

## Abstract

Physical activity (PA) is one of the most fundamental of all traits in the animal kingdom, has pervasive health benefits, and is genetically influenced. Using data from the Million Veteran Program (MVP), we conducted genetic analyses of leisure, work, and home-time PA. For leisure, for individuals of European (EUR) ancestry, n=189,812 and SNP-based heritability (*h*^2^)=0.083±0.005; for African ancestry, n=27,044; *h*^2^= 0.034±0.017; and for Latin-American ancestry, n=10,263; *h*^2^= 0.083±0.036. EUR and cross-ancestry meta-analyses with UK Biobank identified 67 and 70 lead variants. Leisure-time PA was genetically distinct from PA at home or work, with the latter two showing less health benefit on pathologies and lifespan. Mendelian randomization analyses showed protective effects of leisure-time PA on cardiovascular and respiratory system diseases, metabolic traits, aging, and other traits, and there was protective role of leisure-time PA against COVID-19 hospitalization (β=−0.067±0.016; *p*-value=2.8×10^−5^). These findings provide new insights into the biology of PA, showing the causal health benefits of leisure-time PA.

## Introduction

Physical activity (PA) has pervasive and profound health benefits, including beneficial influences on chronic conditions like cancer, hypertension, and type 2 diabetes.^1^ Very different kinds of PA – for example, weightlifting and aerobic exercise - can lead to a longer and healthier life^2^.^2^ An individual’s level of PA is influenced by genetic factors, with estimated heritability (*h*^2^) between 48% and 71% based on twin studies.^3^ To identify genetic variants relevant to PA, genome-wide association studies (GWAS) have been conducted, initially focusing on PA during leisure time.^4–7^ Identifying genetic associations with physical activity affords us a unique opportunity; because we are born with genetic variations prior to exposures to psychological, social, and environmental factors that may affect activity levels, we can begin to decompose some of the complex interactions which underlie and potentially confound the study of this important health trait. There is also a wide range of interindividual differences in requirements for physical activity in daily life. Some jobs require much more physical exertion than others; some individuals must exert more effort in their households than others. These kinds of physical activity tend to differ phenomenologically from exercise taken at leisure, occurring more in concentrated bursts in the first two cases, and with the potential to be more sustained in the latter case; yet level of activity in different contexts is often treated as if equivalent. There are, however, prior reports that comparable health benefits may not be assumed for activity at leisure vs. activity in other contexts^8, 9^, and genetic analyses may help shed light on any differences.

Two powerful previous studies of PA used UK Biobank (UKB) data.^10, 11^ The first study (*N*=91,105) identified three variants associated with PA, based on wrist-worn accelerometer data.^11^ The second study (*N*≤377,234) identified ten significant loci across five phenotypes, three based on self-report data (moderate-to-vigorous PA, vigorous PA, and strenuous sport or other exercise (SSOE) during leisure time), and two on wrist-worn accelerometer data.^10^ A multi-ancestry meta-analysis combining data from 703,901 individuals from 51 studies identified 11 associated loci for moderate-to-vigorous intensity PA.^12^ Combining this phenotype with two traits phenotypically contrary to PA (leisure screen time and sedentary behaviour at work), 99 associated loci were found summing results from PA (11 variants), leisure screen time (89 variants), and sedentary behaviour at work (4 variants); that is, most loci were mapped for phenotypes correlated to a lack of physical activity.^12^

We sought to maximize power to investigate PA genetics by means of a quantitative trait definition, combining different intensities of PA by summing their frequencies, using data from the Million Veteran Program (MVP). We focused on PA *per se*, as opposed to sedentary behavior. Whereas previous studies considered multicontextual PA -- including data from leisure, at work, and home -- our primary quantitative trait, All-PA-Leisure, focused exclusively on PA during leisure time (see Supplementary Methods for PA context definitions). We studied this trait in two meta-analyses (designated “All-PA-Leisure+SSOE”), one of European (EUR) ancestry and one cross-ancestry (EUR, African (AFR), Latin-American (AMR)), with SSOE data from UKB, also based on information of PA during leisure time (Figure S1, Table S1).^10^ This enabled us to investigate, from a genomic perspective, the relationship of PA to a range of health traits. We also investigated PA traits related to the different contexts (home and work, in addition to leisure). In so doing, we greatly increased the number of discovered loci while increasing the diversity of the sample beyond EUR. We investigated protective effects of PA against health traits including type 2 diabetes, gastroesophageal reflux disease, abdominal aortic aneurysm, heart failure, hospitalization caused by COVID-19, osteoarthritis, parental survival, and an aging phenotype. Moreover, we found substantial genetic differences considering PA during the three different contexts (leisure, work, home). These differences are supported by genetic correlation and structural equation modeling analyses.

## Results

### GWAS of All-PA-Leisure (MVP) and All-PA-Leisure+SSOE (MVP+UKB) meta-analysis

#### All-PA-Leisure and SSOE phenotypes

GWAS of All-PA-Leisure in MVP EUR data identified 14 lead SNPs at 13 loci, and GWAS of SSOE in UKB identified 15 lead SNPs in 14 loci (Figure S2-S3, Table S2-S3). The two most significant lead SNPs in the GWAS for All-PA-Leisure (EUR, rs761898, *p*-value=1.63×10^−11^; rs7613360, *p*-value=3.15×10^−11^) map to intergenic regions on chromosome 3 and 14 (Table S3). Additional genome-wide significant loci included *SLC39A8*\*rs13107325 (*p*-value=1.57×10^−10^) (solute carrier family 39 member 8). As previously described,^10^ the most significant variant for GWAS of SSOE in UKB is *CADM2*\*rs62253088 (*p*-value=1.0×10^−19^ in this study) (Cell Adhesion Molecule 2). A sex-stratified analysis of the X chromosome in MVP (All-PA-Leisure) showed no statistically significant associated variants (Figure S4).

#### SNP heritability and genetic correlation of PA phenotypes

In MVP data for All-PA-Leisure, SNP-based heritability (SNP-*h*^2^) is 0.083±0.005 (standard error (SE)) in EUR, 0.034±0.017 in AFR, and 0.083±0.036 in AMR (Table S4). SNP-*h*^2^ of SSOE from UKB is 0.061±0.003. The genetic correlation between All-PA-Leisure and SSOE (r_g_=0.76±0.03; *p*-value=4.7×10^−118^) was higher than the genetic correlation between All-PA-Leisure and the other two traits from the previous UKB report,^10^ moderate-to-vigorous PA (r_g_=0.25±0.04; *p*-value=4.3×10^−11^), and vigorous PA (r_g_=0.55±0.04; *p*-value=3.9×10^−53^). Accordingly, we selected the SSOE phenotype for meta-analyses with MVP data, i.e. the All-PA-Leisure+SSOE meta-analysis (summary statistics results are described in the Supplementary Text, Figure S2-S3, Table S5-S6).

#### All-PA-Leisure+SSOE meta-analyses

The EUR All-PA-Leisure+SSOE meta-analysis greatly increased discovery yielding 67 lead SNPs in 55 loci (Figure 1A, Table S5). The cross-ancestry meta-analysis provided 70 lead SNPs in 56 loci (Figure 1B, Table S6). In both, the strongest association was with *CADM2*\*rs62253088 (EUR meta-analysis *p*-value=2.0×10^−21^; cross-ancestry meta-analysis *p*-value=6.1×10^−20^). The second strongest associated lead SNP was also on chromosome 3: *MST1R*\*rs3733134 (EUR meta-analysis *p*-value=1.4×10^−17^; cross-ancestry meta-analysis *p*-value=6.9×10^−18^).

**Figure 1:**
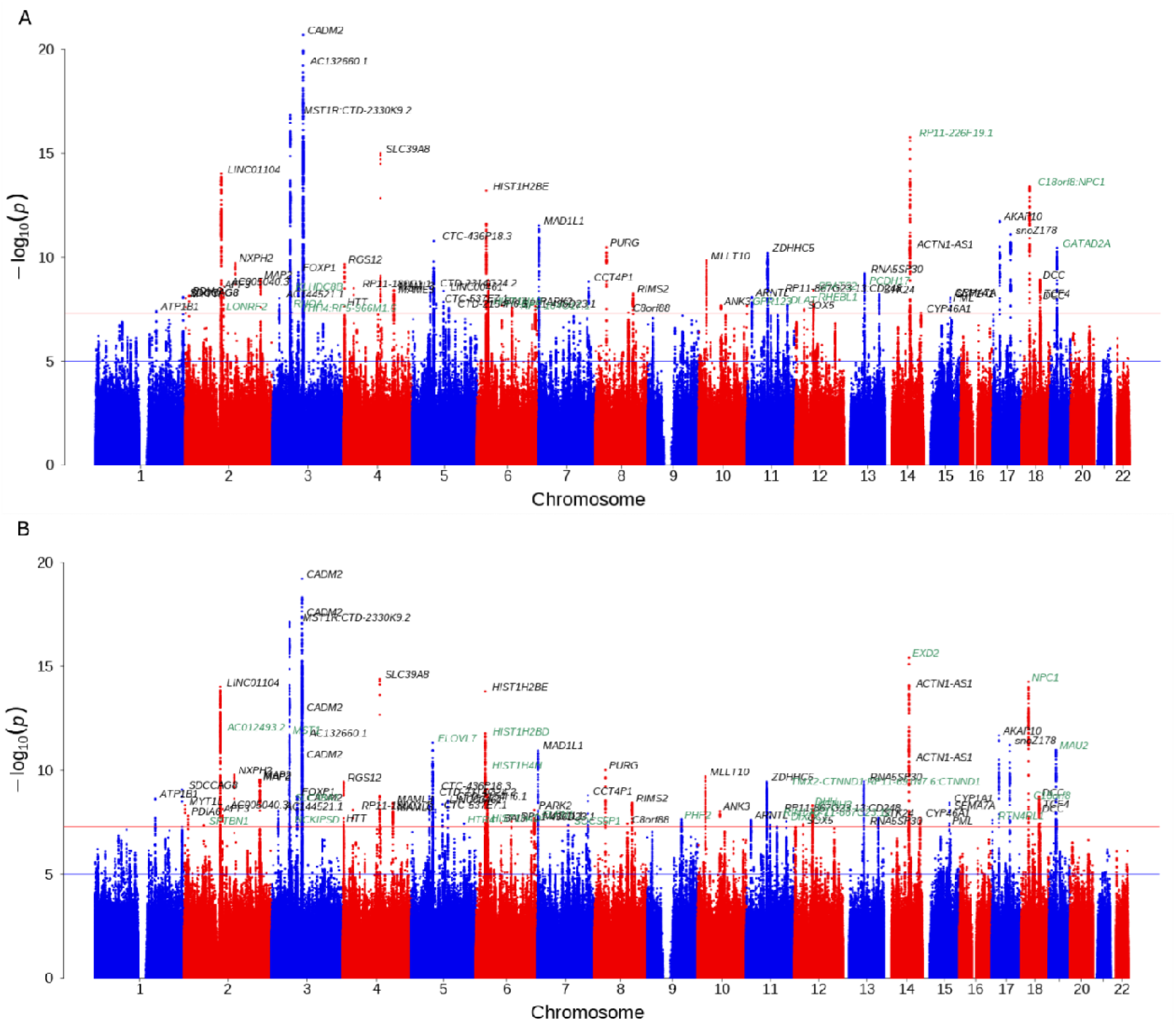
(A) EUR All-PA-Leisure+SSOE meta-analysis and (B) cross-ancestry All-PA-Leisure+SSOE meta-analysis. Annotated genes are the closest to each significant lead SNP. Different lead SNPs are highlighted in green. The red and blue horizontal lines indicate the genome-wide significance (*p*-value=5×10^−8^) and *p*-value=10^−5^ levels, respectively.

#### Gene based, gene-set, tissue expression, cell type enrichment analyses of All-PA-Leisure+SSOE

Focusing on the EUR meta-analysis, the gene-based test using MAGMA identified 119 significant genes from 19,030 protein coding genes (genome-wide significance defined at *p*-value=2.6×10^−6^) (Table S7). *CADM2* was the most significant gene (*p*-value=3.7×10^−25^) followed by *NPC1* (*p*-value=4.8×10^−16^). Four significant gene ontology (GO) terms resulted from the gene-set analysis: structural constituent of the presynapse, presynaptic active zone organization, maintenance of presynaptic active zone structure, and structural constituent of the synapse (Table S8). Tissue Expression Analysis showed 11 significant enrichments in brain tissues, with the two most significant represented in the cerebellar hemisphere and in the cerebellum (Figure S5-S6). For gene-based association analyses and gene-set analyses, similar results were obtained for the cross-ancestry meta-analysis (Supplementary Text, Table S9-S10, Figure S7-S8).

Cell type enrichment analysis was performed with a collection of human cell types from brain, blood, and pancreas tissue (all available tissues). In the EUR meta-analysis of All-PA-Leisure+SSOE, the only significant cell type was *GABA^+^ neurons* from the GSE76381 Linnarsson Human Midbrain dataset (adjusted *p*-value per dataset after multiple testing correction=2.4×10^−5^). In the cross-ancestry meta-analysis, there were no significantly enriched cell types. The results probably reflect the limited number of kinds of tissue included in this analysis.

#### Functional enrichment analysis of All-PA-Leisure+SSOE

Functional enrichment analysis with g:Profiler using significant genes from the gene-based test as input, identified three significant gene ontology terms shared by the EUR meta-analysis and the cross-ancestry meta-analysis (Table S11, S12): presynaptic active zone cytoplasmic component, cell cortex region, and synapse. Also a human phenotype ontology term, polyclonal elevation of IgM, was shared by the EUR meta-analysis and cross-ancestry meta-analysis.

#### PheWAS of All-PA-Leisure+SSOE

Phenome-wide association study (PheWAS) in BioVu revealed significant association with 159 traits (Figure 2, Table S13). The strongest PheWAS results included negative associations with diabetes mellitus (odds ratio (OR)=0.868; *p*-value=5.89×10^−36^), and chronic airway obstruction (OR=0.839; *p*-value=4.22×10^−32^). We also found significant negative associations for traits also analyzed in the genetic correlation analyses: coronary artery disease (ischemic heart disease) (OR=0.892; *p*-value=1.02×10^−21^), gastroesophageal reflux disease (OR=0.929; *p*-value=5.07×10^−13^), cancer of bronchus or lung (OR=0.901; *p*-value=5.47×10^−7^), and asthma (OR=0.937; *p*-value=1.19×10^−5^). We also found a negative correlation with obesity (OR=0.900; *p*-value=2.38×10^− 17^), and several phenotypes related to heart failure: diastolic heart failure (OR=0.880; *p*-value=1.76×10^−9^), systolic or combined heart failure (OR=0.894; *p*-value=1.96×10^−9^), and heart failure (OR=0.902; *p*-value=1.88×10^−5^).

**Figure 2:**
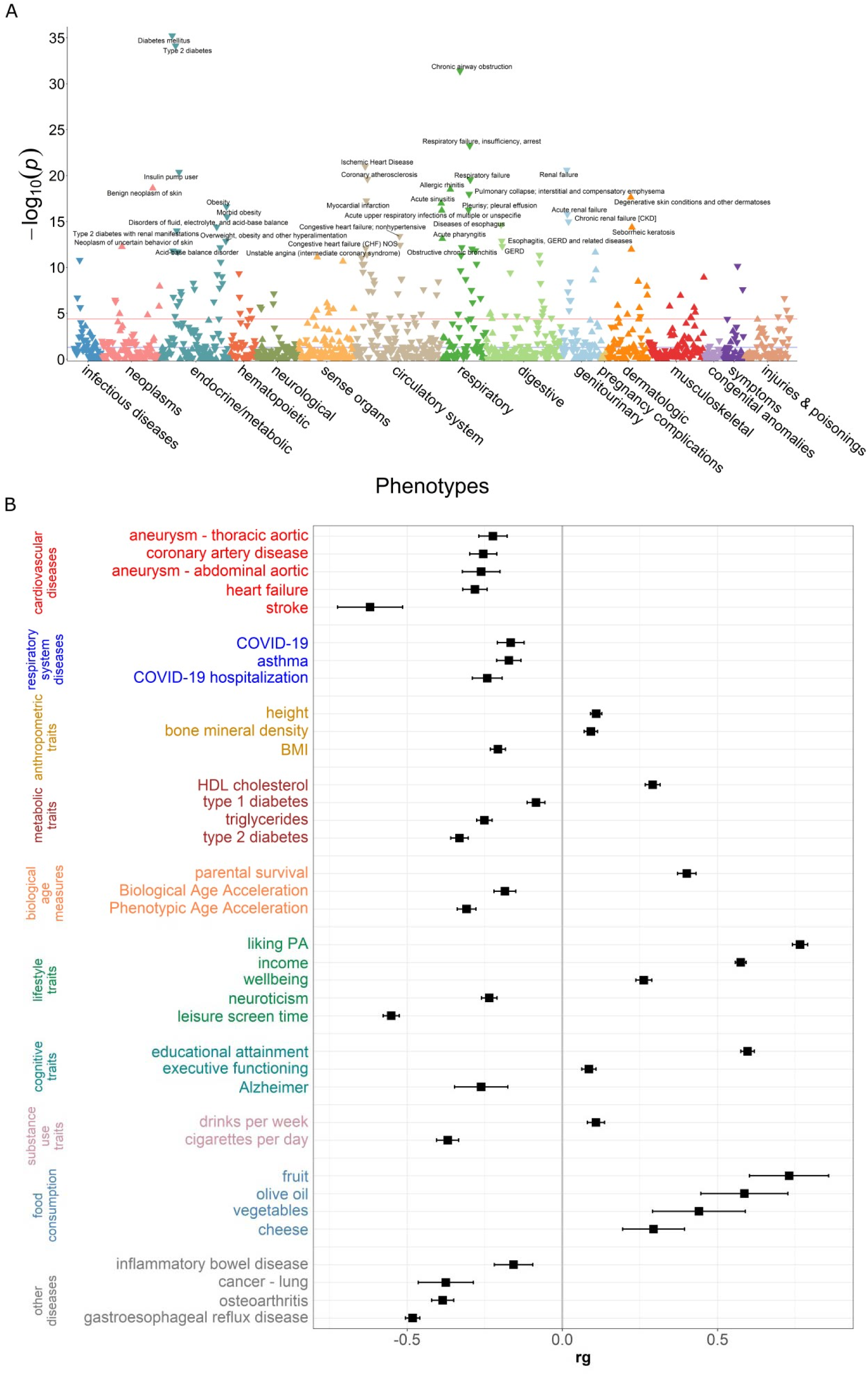
Traits genetically correlated with All-PA-Leisure+SSOE. (A) PheWAS of EUR All-PA-Leisure+SSOE meta-analysis. The red line represents the Bonferroni *p*-value threshold=3.99×10^−5^. (B) Genetic correlations of EUR meta-analysis for All-PA-Leisure+SSOE phenotype with traits of interest. We display here the traits of interest that were significant after the Benjamini-Hochberg false discovery procedure.

#### Genetic correlations: All-PA-Leisure+SSOE and traits of interest

We estimated the genetic correlation of the All-PA-Leisure+SSOE phenotype (EUR meta-analysis) with traits of interest selected to cover a broad range of health diseases and physiological traits, using the Benjamini-Hochberg false discovery rate (FDR) correction (Figure 3, Table S14; more details in *Methods* in Supplementary Text). Results showed significant negative genetic correlations between All-PA-Leisure+SSOE with all disease traits studied, with the strongest values for stroke (r_g_=−0.62±0.11; *p*-value=3.54×10^−9^) and gastroesophageal reflux disease (r_g_=−0.482±0023; *p*-value=9.9×10^−97^) (Table S14 and Supplementary Text).

**Figure 3:**
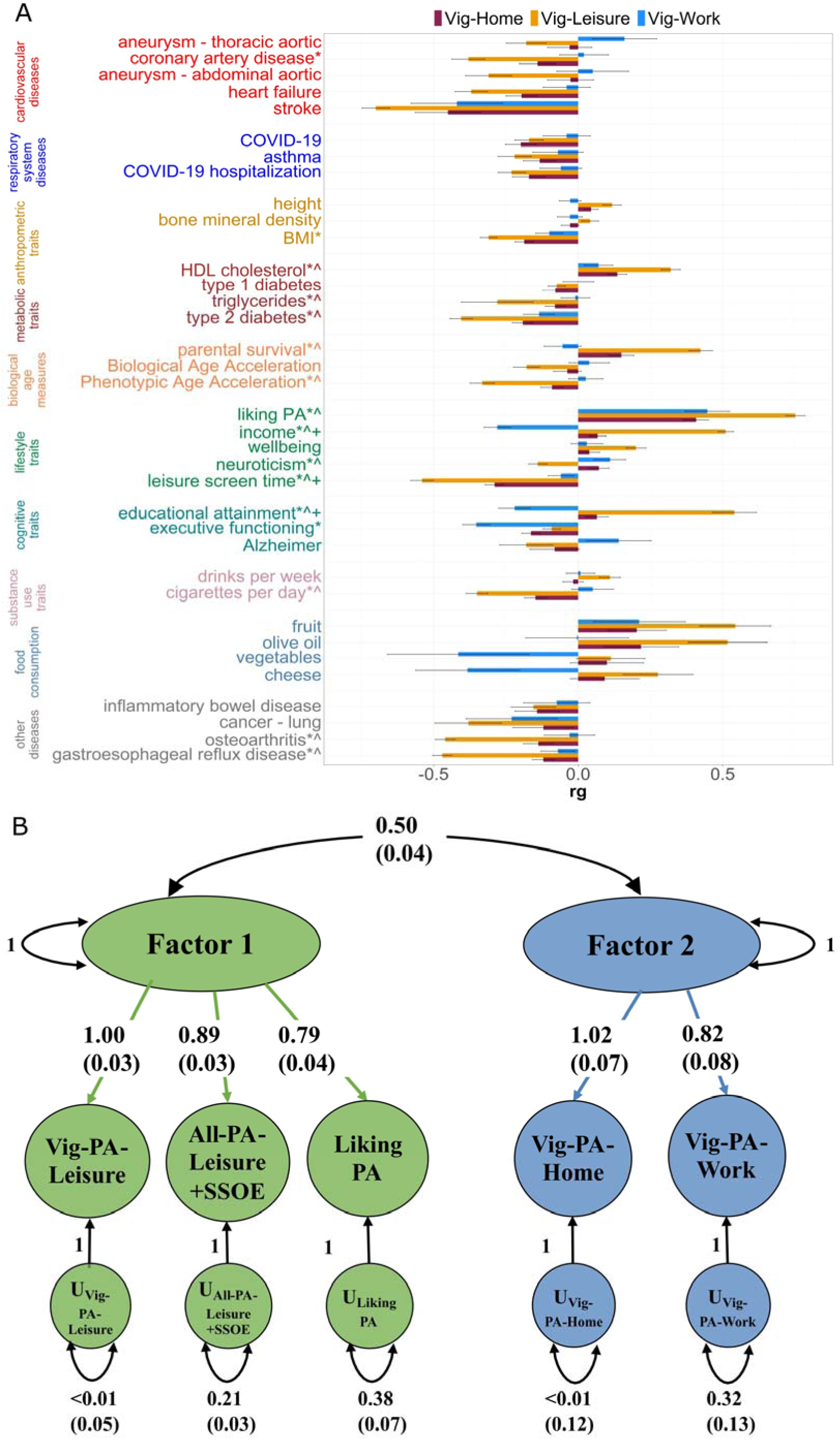
Genetic differences among vigorous PA types. (A) Genetic correlation between each pair of vigorous PA (Vig-PA-Leisure, Vig-PA-Work, Vig-PA-Home) contexts, and other traits of interest. Significantly different correlations were defined as having *p*-value < 4.63×10^−4^ after multiple testing correction: * represents significant difference between Vig-PA-Leisure and Vig-PA-Work, ^ between Vig-PA-Leisure and Vig-PA-Home, + between Vig-PA-Home and Vig-PA-Work. (B) genomic-SEM of the two-factor model, showing the loadings for the inferred traits.

#### Local genetic correlation: All-PA-Leisure+SSOE

We next considered local correlations (LAVA^14^) as global genetic correlations may mask important regional differences. There were a total of 91 significant local bivariate genetic correlations at 61 loci between the EUR All-PA-Leisure+SSOE meta-analysis and the 40 other traits used in the genetic correlation analyses (Table S15). Of these 40 traits, there were 18 with at least one significant local genetic correlation. Seventeen of these traits were previously found to have a significant global genetic correlation with the EUR All-PA-Leisure+SSOE meta-analysis. The trait with the highest number of significant local genetic correlations with All-PA-Leisure+SSOE was educational attainment (31 loci), followed by BMI (15 loci) and income (9 loci). Only one trait-oral cancer, had no significant global genetic correlation with EUR All-PA-Leisure+SSOE but still showed a significant positive local genetic correlation (locus chr11: 118901214-120064528; ρ=0.88; *p*-value=4.5×10^−6^). For two traits, height and wellbeing, we observed local genetic correlations with EUR All-PA-Leisure+SSOE in opposite directions (Table S15).

#### Mendelian Randomization analyses

We used MR analyses to investigate the causal relationship between All-PA-Leisure and traits with which it had significant genetic correlation. Since several statistics for target traits included UKB data, we used the GWAS of All-PA-Leisure in EUR MVP (and not the meta-analysis) to avoid sample overlap. Four methods were tested: MR Egger, Weighted median, Inverse Variance Weighted (IVW), and Simple Mode. We observed significant bidirectional negative (protective) effects of All-PA-Leisure with respect to BMI, type 2 diabetes, leisure screen time, cigarettes per day, and gastroesophageal reflux disease (Tables 1-2). There were unidirectional negative (protective) effects of All-PA-Leisure on abdominal aortic aneurysm, heart failure, COVID-19 hospitalization, triglycerides, Phenotypic Age Acceleration (which is an age-adjusted biological age measure, see Supplementary Methods), and osteoarthritis (Tables 1-2). We found significant bidirectional positive effects between All-PA-Leisure and HDL cholesterol, parental survival, liking of PA, household income, wellbeing, and educational attainment.

**Table 1:**
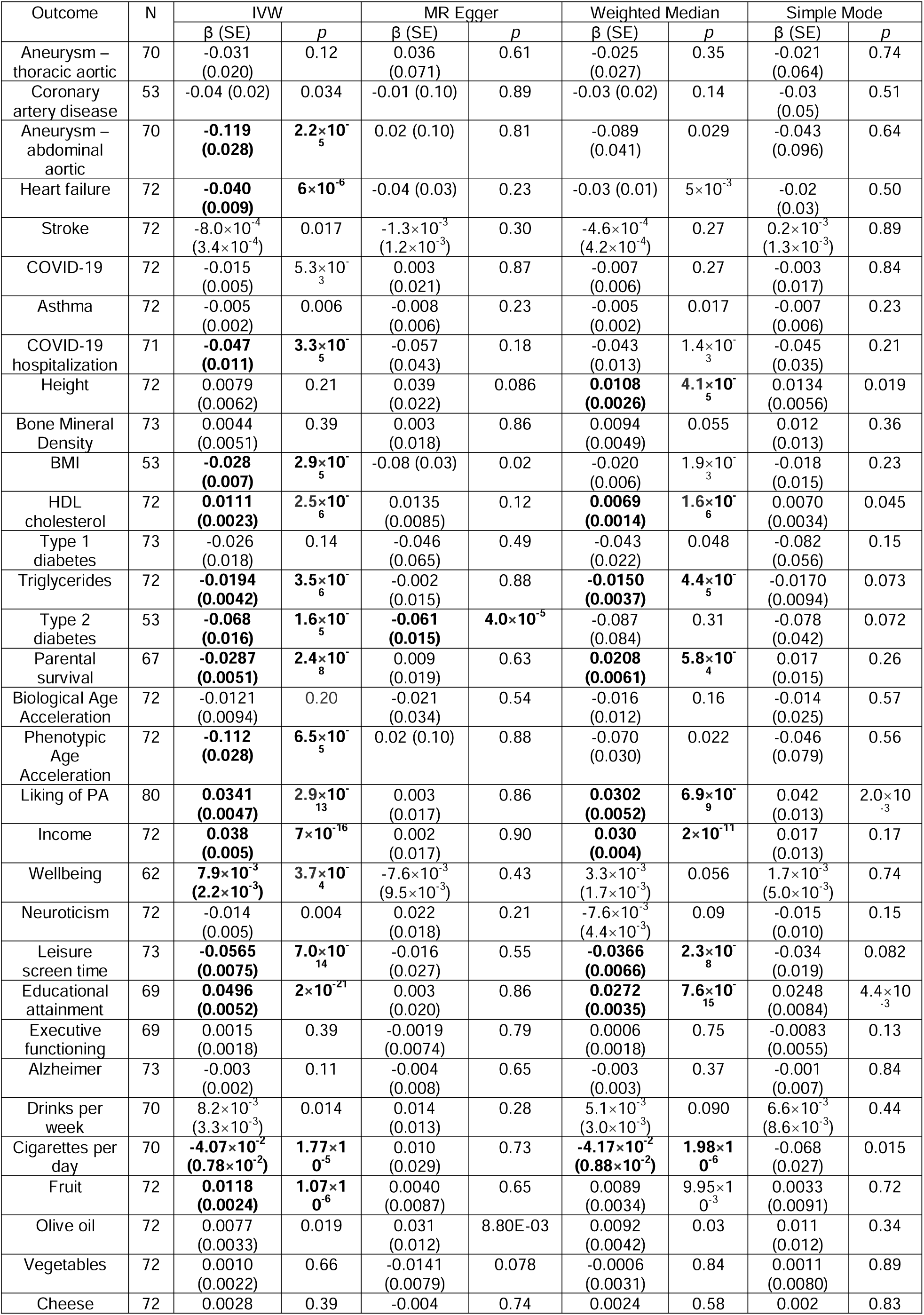
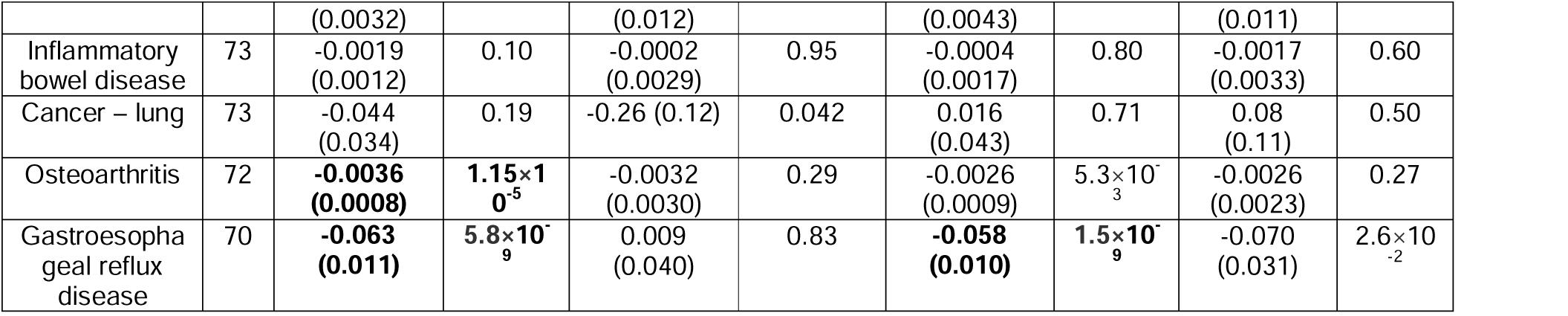
MR analysis of causal effects of PA during leisure time (All-PA-Leisure) on traits of interest. Significant results after multiple testing correction (*p*-value < 6.9×10^−4^) are highlighted in **bold.**

**Table 2:**
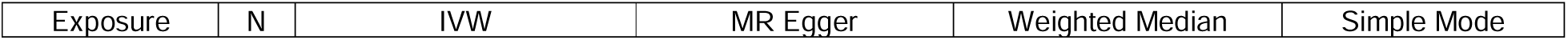
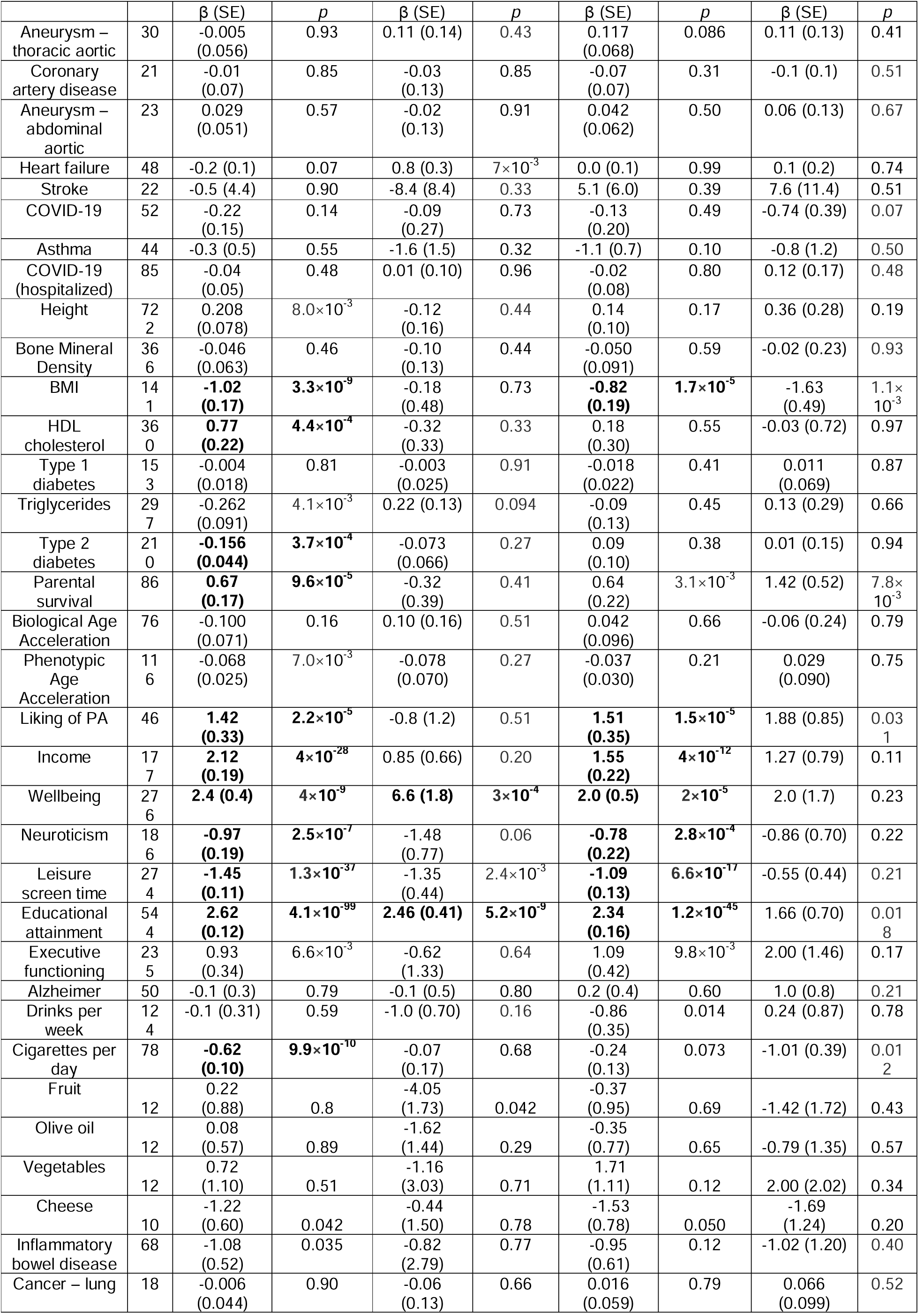
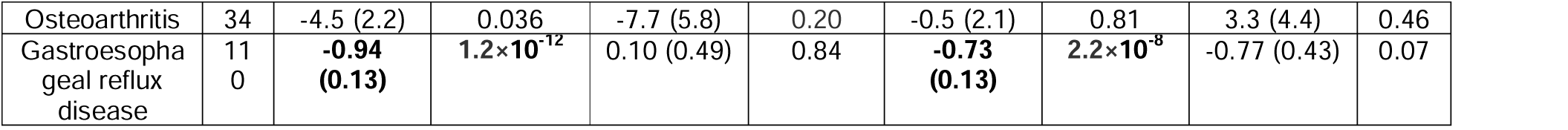
MR analysis of causal effects on PA during leisure time (All-PA-Leisure) of traits of interest. Significant results after multiple testing correction (*p*-value < 6.9×10^−4^) are highlighted in **bold.**

|  |  |  |  |  |  |
| --- | --- | --- | --- | --- | --- |
| Exposure | N | IVW | MR Egger | Weighted Median | Simple Mode |

**Table 2: MR analysis of causal effects on PA during leisure time (All-PA-Leisure) of traits of interest.** Significant results after multiple testing correction ( $p$ -value $< 6.9 \times 10^{-4}$ ) are highlighted in **bold**.
|  |  |  |  |  |  |  |  |  |  |
| --- | --- | --- | --- | --- | --- | --- | --- | --- | --- |
| | | $\beta$ (SE) | $p$ | $\beta$ (SE) | $p$ | $\beta$ (SE) | $p$ | $\beta$ (SE) | $p$ |
| Aneurysm – thoracic aortic | 30 | -0.005 (0.056) | 0.93 | 0.11 (0.14) | 0.43 | 0.117 (0.068) | 0.086 | 0.11 (0.13) | 0.41 |
| Coronary artery disease | 21 | -0.01 (0.07) | 0.85 | -0.03 (0.13) | 0.85 | -0.07 (0.07) | 0.31 | -0.1 (0.1) | 0.51 |
| Aneurysm – abdominal aortic | 23 | 0.029 (0.051) | 0.57 | -0.02 (0.13) | 0.91 | 0.042 (0.062) | 0.50 | 0.06 (0.13) | 0.67 |
| Heart failure | 48 | -0.2 (0.1) | 0.07 | 0.8 (0.3) | $7 \times 10^{-3}$ | 0.0 (0.1) | 0.99 | 0.1 (0.2) | 0.74 |
| Stroke | 22 | -0.5 (4.4) | 0.90 | -8.4 (8.4) | 0.33 | 5.1 (6.0) | 0.39 | 7.6 (11.4) | 0.51 |
| COVID-19 | 52 | -0.22 (0.15) | 0.14 | -0.09 (0.27) | 0.73 | -0.13 (0.20) | 0.49 | -0.74 (0.39) | 0.07 |
| Asthma | 44 | -0.3 (0.5) | 0.55 | -1.6 (1.5) | 0.32 | -1.1 (0.7) | 0.10 | -0.8 (1.2) | 0.50 |
| COVID-19 (hospitalized) | 85 | -0.04 (0.05) | 0.48 | 0.01 (0.10) | 0.96 | -0.02 (0.08) | 0.80 | 0.12 (0.17) | 0.48 |
| Height | 72<br>2 | 0.208 (0.078) | $8.0 \times 10^{-3}$ | -0.12 (0.16) | 0.44 | 0.14 (0.10) | 0.17 | 0.36 (0.28) | 0.19 |
| Bone Mineral Density | 36<br>6 | -0.046 (0.063) | 0.46 | -0.10 (0.13) | 0.44 | -0.050 (0.091) | 0.59 | -0.02 (0.23) | 0.93 |
| BMI | 14<br>1 | **-1.02 (0.17)** | **$3.3 \times 10^{-9}$** | -0.18 (0.48) | 0.73 | **-0.82 (0.19)** | **$1.7 \times 10^{-5}$** | -1.63 (0.49) | $1.1 \times 10^{-3}$ |
| HDL cholesterol | 36<br>0 | **0.77 (0.22)** | **$4.4 \times 10^{-4}$** | -0.32 (0.33) | 0.33 | 0.18 (0.30) | 0.55 | -0.03 (0.72) | 0.97 |
| Type 1 diabetes | 15<br>3 | -0.004 (0.018) | 0.81 | -0.003 (0.025) | 0.91 | -0.018 (0.022) | 0.41 | 0.011 (0.069) | 0.87 |
| Triglycerides | 29<br>7 | -0.262 (0.091) | $4.1 \times 10^{-3}$ | 0.22 (0.13) | 0.094 | -0.09 (0.13) | 0.45 | 0.13 (0.29) | 0.66 |
| Type 2 diabetes | 21<br>0 | **-0.156 (0.044)** | **$3.7 \times 10^{-4}$** | -0.073 (0.066) | 0.27 | 0.09 (0.10) | 0.38 | 0.01 (0.15) | 0.94 |
| Parental survival | 86 | **0.67 (0.17)** | **$9.6 \times 10^{-5}$** | -0.32 (0.39) | 0.41 | 0.64 (0.22) | $3.1 \times 10^{-3}$ | 1.42 (0.52) | $7.8 \times 10^{-3}$ |
| Biological Age Acceleration | 76 | -0.100 (0.071) | 0.16 | 0.10 (0.16) | 0.51 | 0.042 (0.096) | 0.66 | -0.06 (0.24) | 0.79 |
| Phenotypic Age Acceleration | 11<br>6 | -0.068 (0.025) | $7.0 \times 10^{-3}$ | -0.078 (0.070) | 0.27 | -0.037 (0.030) | 0.21 | 0.029 (0.090) | 0.75 |
| Liking of PA | 46 | **1.42 (0.33)** | **$2.2 \times 10^{-5}$** | -0.8 (1.2) | 0.51 | **1.51 (0.35)** | **$1.5 \times 10^{-5}$** | 1.88 (0.85) | 0.03<br>1 |
| Income | 17<br>7 | **2.12 (0.19)** | **$4 \times 10^{-28}$** | 0.85 (0.66) | 0.20 | **1.55 (0.22)** | **$4 \times 10^{-12}$** | 1.27 (0.79) | 0.11 |
| Wellbeing | 27<br>6 | **2.4 (0.4)** | **$4 \times 10^{-9}$** | **6.6 (1.8)** | **$3 \times 10^{-4}$** | **2.0 (0.5)** | **$2 \times 10^{-5}$** | 2.0 (1.7) | 0.23 |
| Neuroticism | 18<br>6 | **-0.97 (0.19)** | **$2.5 \times 10^{-7}$** | -1.48 (0.77) | 0.06 | **-0.78 (0.22)** | **$2.8 \times 10^{-4}$** | -0.86 (0.70) | 0.22 |
| Leisure screen time | 27<br>4 | **-1.45 (0.11)** | **$1.3 \times 10^{-37}$** | -1.35 (0.44) | $2.4 \times 10^{-3}$ | **-1.09 (0.13)** | **$6.6 \times 10^{-17}$** | -0.55 (0.44) | 0.21 |
| Educational attainment | 54<br>4 | **2.62 (0.12)** | **$4.1 \times 10^{-99}$** | **2.46 (0.41)** | **$5.2 \times 10^{-9}$** | **2.34 (0.16)** | **$1.2 \times 10^{-45}$** | 1.66 (0.70) | 0.01<br>8 |
| Executive functioning | 23<br>5 | 0.93 (0.34) | $6.6 \times 10^{-3}$ | -0.62 (1.33) | 0.64 | 1.09 (0.42) | $9.8 \times 10^{-3}$ | 2.00 (1.46) | 0.17 |
| Alzheimer | 50 | -0.1 (0.3) | 0.79 | -0.1 (0.5) | 0.80 | 0.2 (0.4) | 0.60 | 1.0 (0.8) | 0.21 |
| Drinks per week | 12<br>4 | -0.1 (0.31) | 0.59 | -1.0 (0.70) | 0.16 | -0.86 (0.35) | 0.014 | 0.24 (0.87) | 0.78 |
| Cigarettes per day | 78 | **-0.62 (0.10)** | **$9.9 \times 10^{-10}$** | -0.07 (0.17) | 0.68 | -0.24 (0.13) | 0.073 | -1.01 (0.39) | 0.01<br>2 |
| Fruit | 12 | 0.22 (0.88) | 0.8 | -4.05 (1.73) | 0.042 | -0.37 (0.95) | 0.69 | -1.42 (1.72) | 0.43 |
| Olive oil | 12 | 0.08 (0.57) | 0.89 | -1.62 (1.44) | 0.29 | -0.35 (0.77) | 0.65 | -0.79 (1.35) | 0.57 |
| Vegetables | 12 | 0.72 (1.10) | 0.51 | -1.16 (3.03) | 0.71 | 1.71 (1.11) | 0.12 | 2.00 (2.02) | 0.34 |
| Cheese | 10 | -1.22 (0.60) | 0.042 | -0.44 (1.50) | 0.78 | -1.53 (0.78) | 0.050 | -1.69 (1.24) | 0.20 |
| Inflammatory bowel disease | 68 | -1.08 (0.52) | 0.035 | -0.82 (2.79) | 0.77 | -0.95 (0.61) | 0.12 | -1.02 (1.20) | 0.40 |
| Cancer – lung | 18 | -0.006 (0.044) | 0.90 | -0.06 (0.13) | 0.66 | 0.016 (0.059) | 0.79 | 0.066 (0.099) | 0.52 |
| Osteoarthritis | 34 | -4.5 (2.2) | 0.036 | -7.7 (5.8) | 0.20 | -0.5 (2.1) | 0.81 | 3.3 (4.4) | 0.46 |
| Gastroesophageal reflux disease | 110 | **-0.94 (0.13)** | **$1.2 \times 10^{-12}$** | 0.10 (0.49) | 0.84 | **-0.73 (0.13)** | **$2.2 \times 10^{-8}$** | -0.77 (0.43) | 0.07 |

#### MVMR analyses testing BMI as confounder

To investigate the protective effect of All-PA-Leisure with respect to COVID-19 further, we considered a possible role of BMI as confounder, as high BMI leads to worse COVID-19 outcomes.^15^ We performed a multivariable MR (MVMR), where we used the All-PA-Leisure phenotype and BMI as exposures, and COVID-19 hospitalization as the outcome. We observed a significant protective effect of All-PA-Leisure on the COVID-19 trait which considers hospitalized cases (effect=−0.067±0.016; *p*-value=2.8×10^−5^; Table 3). Considering the role that BMI plays for many other health outcomes, we also conducted MVMR including BMI for all other traits that had significant MR results. Despite the reduced power of the analysis due to a decrease in number of instrumental variables, which were reduced because of the required SNP overlap needed between three GWAS compared to the two GWAS for the traditional MR analysis, we found significant protective effects of All-PA-Leisure on HDL cholesterol, triglycerides, osteoarthritis, and gastroesophageal reflux disease (Table 3).

**Table 3:** MVMR analysis of All-PA-Leisure setting BMI as exposures. Significant results after multiple testing correction (*p*-value < 5.6×10^−3^ outcomes) are highlighted in **bold.**

| Exposure | Outcome | N | $\beta$ (SE) | $p$ |
| --- | --- | --- | --- | --- |
| PA | Aneurysm – abdominal aortic | 7 | -0.087 (0.055) | 0.11 |
| BMI | Aneurysm – abdominal aortic | 49 | 0.31 (0.18) | 0.092 |
| **PA** | **COVID-19 hospitalization** | **7** | **-0.067 (0.016)** | **$2.8 \times 10^{-5}$** |
| BMI | COVID-19 hospitalization | 50 | 0.37 (0.05) | $8.2 \times 10^{-13}$ |
| **PA** | **HDL cholesterol** | **7** | **0.0221 (0.0047)** | **$2.3 \times 10^{-6}$** |
| BMI | HDL cholesterol | 50 | -0.042 (0.016) | $7.0 \times 10^{-3}$ |
| **PA** | **Triglycerides** | **7** | **-0.038 (0.011)** | **$3.6 \times 10^{-4}$** |
| BMI | Triglycerides | 50 | 0.040 (0.035) | 0.26 |
| PA | Type 2 diabetes | 6 | -0.058 (0.057) | 0.30 |
| BMI | Type 2 diabetes | 43 | 0.42 (0.18) | 0.19 |
| PA | Parental survival | 7 | 0.030 (0.010) | $2.9 \times 10^{-3}$ |
| BMI | Parental survival | 48 | -0.171 (0.034) | $4.4 \times 10^{-7}$ |
| PA | Phenotypic Age Acceleration | 7 | -0.091 (0.044) | $3.7 \times 10^{-2}$ |
| BMI | Phenotypic Age Acceleration | 50 | 0.92 (0.15) | $4.8 \times 10^{-10}$ |
| **PA** | **Osteoarthritis** | **7** | **-0.0037 (0.0012)** | **$1.4 \times 10^{-3}$** |
| BMI | Osteoarthritis | 50 | 0.0253 (0.0039) | $1.3 \times 10^{-10}$ |
| **PA** | **Gastroesophageal reflux disease** | **7** | **-0.063 (0.015)** | **$2.3 \times 10^{-5}$** |
| BMI | Gastroesophageal reflux disease | 50 | 0.141 (0.050) | $4.6 \times 10^{-3}$ |

#### All-PA-Leisure&Home and All-PA-Leisure&Home&Work phenotypes

To evaluate possible gain of information from combining data from PA during leisure time with PA performed during home and work time, we ran two more GWAS in EUR. First, we summed All-PA-Leisure with All-PA from physical activity at home (All-PA-Leisure&Home; Supplementary Methods, Figure S9). We next added information from All-PA at work (All-PA-Leisure&Home&Work; Supplementary Methods, Figure S10). In both the new GWASs we observed a substantial reduction of the number of association peaks compared to All-PA-Leisure, as would be expected given the relatively low genetic correlations among these traits.

For the EUR All-PA-Leisure+SSOE meta-analysis, we also conducted TWAS, fine-mapping, and MTAG analysis with leisure screen time and liking of PA (see Supplementary Text, Table S16-S20).

### GWAS of Vigorous PA phenotypes (Leisure, at Home, at Work)

#### Kinds of vigorous physical activity and their genetic interrelationships

We ran independent MVP GWAS of all vigorous activity types depending by the context (Vig-PA-Leisure, Vig-PA-Home, Vig-PA-Work) in EUR, and investigated correlations between each set of two. For Vig-PA-Leisure and Vig-PA-Home, r_g_=0.64±0.04 (*p*-value=5.0×10^−59^); Vig-PA-Leisure and Vig-PA-Work, r_g_=0.46±0.07 (*p*-value=9.8×10^−11^). Vig-PA-Home and Vig-PA-Work were more strongly correlated with each other (r_g_=0.97±0.05; *p*-value=1.2×10^−73^). SNP-*h*^2^ of vigorous PA phenotypes are reported in Table S21.

For each one of the three vigorous PA traits, we ran GWAS, and for Vig-PA-Leisure we also conducted XWAS, gene-based testing, TWAS, fine-mapping, functional enrichment, mitochondria-SNV association, and gene-based association analysis on the mitochondrial genome (see Supplementary Text, Table S22-S28, Figure S11-S15).

#### Genetic correlations between vigorous PA traits and traits of interest

To understand the differences between the vigorous activity traits better, we calculated the genetic correlations between Vig-PA-Leisure, Vig-PA-Work, and Vig-PA-Home individually with the previously analyzed traits of interest. We observed more statistically-significant differences between Vig-PA-Leisure and Vig-PA-Work (for 15 traits), than when considering Vig-PA-Leisure and Vig-PA-Home (for 12 traits) or Vig-PA-Home and Vig-PA-Work (for 3 traits) (Figure 4a). Among these three pairs of vigorous activity traits, there were numerous significant differences regarding the genetic correlations with income, leisure screen time, and educational attainment. Analyses performed adjusting the vigorous PA traits with income using mtCOJO are described in Supplementary Results, Table S29, Figure S16.

#### Genomic-SEM analysis

Considering the differences identified among the three vigorous PA traits (e.g. Vig-PA-Leisure, Vig-PA-Work, and Vig-PA-Home), we used genomic-SEM analysis^16^ to understand their relationship better, inferring the overall genetic architecture among them and two other physical activity traits in EUR, All-PA-Leisure+SSOE and liking of PA. The exploratory factor analysis (EFA) indicated that a two-factor model fit the data best (Figure 4b). The two-factor model accounted for 91% of cumulative variance explained (Factor 1: 50%; Factor 2: 41%). Both factors had strong strength of sum of squares (SS) loadings (Factor 1 SS = 2.50; Factor 2 SS = 2.04). Confirmatory factor analysis (CFA) confirmed that a two correlated-factors model fit the data well with a comparative fit index (CFI) of 0.97, chi-square=109.03, Aikake information criterion (AIC)=129.03, and a standardized root mean square residual (SRMR) of 0.069. Vig-PA-Leisure (loading=1.00±0.03), All-PA-Leisure+SSOE (loading=0.89±0.03), and liking of PA (loading=0.79±0.04) loaded most strongly on Factor 1, while Vig-PA-Home (loading=1.02±0.07) and Vig-PA-Work (loading=0.82±0.08) loaded most strongly on Factor 2 (Figure 4b). Factor 1 and Factor 2 were correlated at 0.50 (0.04).

#### Survival analyses

Survival analyses showed a significant reduction of risk of all-cause death for Vig-PA-Leisure (beta=−0.1173±0.0037; *p*-value≤2×10^−16^) and for Vig-PA-Work (beta=−0.0061±0.0029; *p*-value=0.036). There was no statistically significant reduction of risk of death for Vig-PA-Home, though the direction of effect and effect size were similar (beta=−0.0052±0.0030; *p*-value=0.082).

## Discussion

Physical activity (PA) has long been appreciated to ameliorate risk for illness. Increasing physical activity on a population level could greatly reduce morbidity and mortality – this is a major public health undertaking and very difficult to accomplish. Understanding genetic factors that influence level of activity for individuals could provide pathways for medical interventions to support efforts to increase PA. We present here comprehensive information on the genetics of PA and investigate the relationships between PA and health. This work provides tools to dissect causal relationships between a range of other traits and PA, and to uncover previously unknown biology responsible in part for inter-individual differences in activity. Those relationships were remarkably different depending on the context of the physical activity – in particular, whether the activity was associated with PA during leisure time (protective) rather than at work or around the house (often not protective). Our main focus was accordingly on leisure-PA. All-PA-Leisure considered different levels of PA intensity and frequency. We completed GWAS of All-PA-Leisure for EUR, AFR, and AMR ancestry in MVP. We combined these data with UKB SSOE trait in two meta-analyses, one EUR ancestry and the other cross-ancestry.

The polygenic architecture of leisure-time PA showed genetic liability to PA as a protective effect with respect to numerous diseases. It was previously reported that PA has negative genetic correlation with type 2 diabetes^10, 12, 17^, triglyceride level^10^, rheumatoid arthritis^12^, coronary artery disease^12^, and lung cancer^12^. There is prior genetic evidence of the role of PA on preventing colorectal cancer using MR methods^18^, but we did not find a significant genetic correlation with colorectal cancer. Our analyses confirmed the negative genetic correlations with type 2 diabetes, coronary artery disease, and lung cancer. Moreover, we found significant negative genetic correlation between PA and other medically relevant traits: thoracic and abdominal aortic aneurysm, heart failure, stroke, asthma, type 1 diabetes, Alzheimer’s disease, inflammatory bowel disease, osteoarthritis, and gastroesophageal reflux disease. Notably, PA was also protective with respect to COVID-19 and COVID-19-associated hospitalization, and positively correlated with parental survival.

Whereas genetic correlations are an average of the shared association across the genome, local genetic correlations can detect regions of shared genetic associations, including finding signals in opposing directions that a genome-wide analysis would not be able to capture. Local genetic correlation, like global correlation, supported a protective role of PA for multiple health traits, showing negative local genetic correlations for diabetes types 1 and 2, gastroesophageal reflux disease, and osteoarthritis. We also found negative local genetic correlations between All-PA-Leisure+SSOE and BMI, triglycerides, Phenotypic Age Acceleration, leisure screen time, neuroticism, and cigarettes per day. Positive local genetic correlations were found between All-PA-Leisure+SSOE and bone mineral density, income, liking of PA, educational attainment, and, unexpectedly, oral cancer: there was a single locus showing positive local genetic correlation between PA and oral cancer – however, genome-wide genetic correlation between these traits calculated with LDSC was not significant (*p*-value=0.418). We also found local genetic correlations in opposite directions – at different genomic locations – with All-PA-Leisure+SSOE for height and wellbeing.

Our MR results confirmed a protective effect of All-PA-Leisure genetic liability against type 2 diabetes, abdominal aortic aneurysm, heart failure, COVID-19 associated hospitalization, triglyceride level, osteoarthritis, and gastroesophageal reflux disease. Most of these causal effects were seen even when we tested for BMI as a confounder in the MR (MVMR) results (with the exception of heart failure, which we could not test because of sample overlap for the BMI and heart failure datasets). The low number of instrumental variables in our MVMR analyses could result in false negatives. These findings nevertheless confirm the utility of All-PA-Leisure to decrease risk for numerous diseases.^19, 20^ MR results also suggested a negative effect of All-PA-Leisure versus the biological age measure Phenotypic Age Acceleration, and a bidirectional effect between All-PA-Leisure and parental survival, uncovering the protective effect of genetic liability for this PA phenotype versus aging and mortality.

PheWAS analysis in BioVU^21^, which allowed a broad exploration of disease traits in an independent sample, showed negative correlation with PA with several previously mentioned health traits including diabetes mellitus, cardiovascular disease (e.g. coronary artery disease and heart failure), gastroesophageal reflux disease, lung cancer, and asthma. The strongest association was related to diabetes mellitus (*p*-value=5.89×10^−36^).

The EUR meta-analysis of All-PA-Leisure+SSOE identified 163 independent significant SNPs, 91 of them novel compared to the independent significant SNPs of the two GWASs, All-PA-Leisure and SSOE, in the meta-analysis. *CADM2*\*rs62253088 (encoding Cell Adhesion Molecule 2), on chromosome 3, had the strongest association in both EUR and cross-ancestry All-PA-Leisure+SSOE meta-analyses (EUR meta-analysis *p*-value=2.0×10^−21^; cross-ancestry (EUR, AFR, AMR) meta-analysis *p*-value=6.1×10^−20^), but it was not significant in the GWAS of Vig-PA-Leisure. However, considering Vig-PA-Leisure, we found another variant mapping to *CADM2* which was a significant lead SNP in the GWAS (rs2326266; *p*-value=4.1×10^−8^) and *CADM2* also had the most significant *p*-value in the gene-based test (*p*-value=2.5×10^−8^). *CADM2* variants, including rs62253088, have previously been associated to PA, and also to decreased neuroticism and decreased self-reported nervous and anxious feelings.^10^ *CADM2* was also previously found to be associated to obesity and metabolic traits,^22^ cannabis use,^23^ smoking and alcohol related traits,^24^ risk-taking behaviour,^25^ educational attainment,^26^ and autism spectrum disorder.^27^ *CADM2* encodes a member of the synaptic cell adhesion molecule 1 (SynCAM) family which belongs to the (Ig) immunoglobulin superfamily.

The second most significant variant of the two meta-analyses, *MST1R*\*rs3733134 (EUR meta-analysis *p*-value=1.4×10^−17^; cross-ancestry meta-analysis *p*-value=6.9×10^−18^), encoding macrophage stimulating 1 receptor, was also significant in the GWAS of Vig-PA-Leisure (*p*-value=2.5×10^−8^). The third most significant variant was *EXD2*\*rs4899292 (EUR meta-analysis *p*-value=1.7×10^−16^; cross-ancestry meta-analysis *p*-value=3.8×10^−16^). *EXD2* (encoding exonuclease 3’-5’ domain containing 2) is involved in DNA replication to stabilize and restart stalled replication forks.^28^ Other strong significant associations included genes *LONRF2* (chromosome 2; encoding LON peptidase N-terminal domain and ring finger 2) and *SLC39A8*, involved in the binding activity and transportation of metal ions, and *NPC1* (chromosome 18; encoding NPC intracellular cholesterol transporter 1), involved in cholesterol transport. *SLC39A8* was previously associated to several psychiatric disorders, including problematic alcohol use,^29^ schizophrenia,^30^ and as a lead SNP in opioid use disorder.^31^ *C18orf8*, also known as *RMC1* (chromosome 18; encoding regulator of MON1-CCZ1), mapped to the second most significant peak in the gene-based test of the cross-ancestry meta-analysis, and is associated with type 2 diabetes and BMI.^32–34^

Post-GWAS analyses demonstrated that PA is related to brain structure and cellular architecture. From MAGMA tissue expression analyses we determined that cerebellum and cerebellar hemisphere showed significant enrichments. Cerebellar function is critical for skills relevant to PA, such as coordination and movement. These results were similar to results of the previous GWAS of SSOE.^10^ Cell type specificity analyses showed a significant association of PA with the GABA^+^ neurons.^35^ EUR All-PA-Leisure+SSOE and cross-ancestry meta-analyses shared three significant gene sets in their MAGMA gene-set analysis results: structural constituent of presynapse, presynaptic active zone organization, and maintenance of presynaptic active zone structure. This finding highlights the role of presynaptic cells in PA and the importance of brain in the biology of PA level.

Vig-PA-Leisure was, as noted, genetically different from Vig-PA-Home and at Vig-PA-Work. We observed moderate genetic correlation between Vig-PA-Leisure and Vig-PA-Home (r_g_=0.64±0.04; *p*-value=5.0×10^−59^), but lower genetic correlation between Vig-PA-Leisure and Vig-PA-Work (r_g_=0.46±0.07; *p*-value=9.8×10^−11^). While there was high genetic correlation between Vig-PA-Home and Vig-PA-Work (r_g_=0.97±0.05; *p*-value=1.2×10^−73^), there were differences between these two traits in genetic correlation analyses with other traits of interest, in particular for income, educational attainment, and leisure screen time. Previous work on dementia also showed genetic differences between PA during leisure vs. work time ^36^.

Consistent with what we observed with the divergent patterns of genetic correlation, the genomic-SEM results supported that physical activity traits are genetically heterogeneous. Traits more similar to voluntary physical activity and healthy lifestyle habits loaded on Factor 1 while Factor 2 captured traits more consistent with strenuous work and labor performed as part of work duties and responsibilities at home. When we adjusted the vigorous physical activity traits for income using mtCOJO, we still observed differences on genetic correlations with health traits. These findings suggest that not all contexts of physical activity are equally beneficial, and some forms of vigorous activity may be more strongly linked to improved mental and physical health.

A further confirmation of the different benefits by PA context was provided by the survival analyses, which show a strong significant reduction of risk of death for Vig-PA-Leisure (beta=−0.1173±0.0037; *p*-value≤2×10^−16^) compared to a weak effect for Vig-PA-Work (beta=−0.0061±0.0029; *p*-value=0.036), and none for for Vig-PA-Home (beta=−0.0052±0.0030; *p*-value=0.082).The small protective effect for Vig-PA-Work contrasts with the opposing directions for risk for several other important health traits (Figure 4a). The survival analyses may be confounded by health status (i.e., healthier individuals are able to exercise more vigorously) and competing risks (death). We conclude that PA at leisure is the most protective with respect to negative medical outcomes.

This study has several limitations, including a relative lack of females in the MVP dataset (approximately 9% for our data which nevertheless equates to >17,000 females for Vig-PA-Leisure), which also made the potential interpretation of sex-specific associations infeasible due to power imbalance. UKB has excellent female representation. Moreover, while there are many non-EUR subjects included, their numbers are still insufficient, such that the cross-ancestry analysis mostly reflects findings in EUR. It would also be important to verify the tissue expression analyses, which are currently highlighting only significant associations with the brain, with more tissues and larger samples. Indeed, our results may reflect a lack of power for other tissues or our sample size -- of the 53 tissue types contained in Genotype-Tissue Expression version 8 (GTEx v8), 13 tissues are brain specific.

In conclusion, we have made progress towards revealing the genetic and biological nature of PA during leisure time, which can differ greatly from PA undertaken in other contexts. In effect, physical activity that one chooses to engage in in a leisure context has many positive effects with respect to health traits, while activity that one must do, in a work or a household context, tends not to have these beneficial effects. This difference was still observed after we adjusted for income. Moreover, PA in the leisure context shows a much stronger decrease in the risk of death compared to the other contexts. This may reflect the differences in intensity and duration of leisure compared with work and home activity in the industrialized era. We detected a protective role of PA genetic liability on several diseases, using global and local genetic correlation, PheWAS analyses and, importantly, causal inference using MR. MR analyses showed a significant negative effect of PA on several traits, with significant protective relationships on type 2 diabetes, abdominal aortic aneurysm, heart failure, triglyceride level, osteoarthritis, gastroesophageal reflux disease, and COVID-19 associated hospitalization. Finally, these results provide genetic insights into the biological bases of PA, and add to strong experimental, epidemiological, and observational literature showing the value to health of physical activity.

## Data availability

All MVP summary statistics are made available through dbGAP request under accession phs001672.v7.p1.

Meta-analysis summary statistics are available through the Gelernter lab website: https://medicine.yale.edu/lab/gelernter/.

## Acknowledgements

This research is based in part on data from the Million Veteran Program, Office of Research and Development, Veterans Health Administration.

This work was supported by funding from the Department of Veterans Affairs Office of Research and Development, USVA, grants I01CX001849, and the VA Cooperative Studies Program study, no. 575B; the VA National Center for PTSD Research, and the West Haven VA Mental Illness

Research, Education and Clinical Center; and by NIH grants R01DA054869 (JG), R01 AA026364 (JG). D.F.L. is supported by a Career Development Award CDA-2 from the Veterans Affairs Office of Research and Development (1IK2BX005058-01A2) and is Aimee Mann Fellow of Psychiatric Genetics.

This publication does not represent the views of the Department of Veteran Affairs or the United States Government.

