## Supplemental Material for "A contextual genomic perspective on physical activity and its relationship to health, well-being and illness"

Supplementary Text

### Methods

**MVP datasets**

The U.S. Department of Veterans Affairs (VA) Million Veteran Program (MVP) is one of the largest and most diverse biobanks in the world with genetic and electronic health record (EHR) data available.^1,2^ Ethical approval of the MVP study was given by The Central VA Institutional Review Board (IRB) and site-specific IRBs. All relevant ethical regulations for work with human subjects were followed in the conduct of the study, and informed consent was obtained from all participants.

Participants provided a blood sample for genomic analyses, granted access to medical records, and agreed to complete two questionnaires, the MVP Baseline and Lifestyle Surveys. PA information used in this study was collected from the MVP Lifestyle Survey, and data were subdivided according to context (leisure, work, and home), intensity (vigorous, moderate, and light), and frequency (Figure S1). The three levels of activity were defined as following:

1) *Vigorous*: activities that cause your heart to beat rapidly and you work up a good sweat and are breathing heavily; performed at least 10 minutes at a time;

2) *Moderate*: activities that cause your heart rate to increase slightly and you typically work up a sweat, but are not physically exhausting; performed for at least 10 minutes at a time;

3) *Light*: activities that require little physical effort”.

The levels of activity were considered together with frequency information, which could be “daily”, “several times/week”, “once/week”, “several times/month”, “once/month or less”, or “never”, when participants answered to three questions regarding the place where PA was effected. The questions asked to participants in the MVP Lifestyle Survey were the following:

During the job time: *while at your job, how often do you engage in the following levels of activity?*

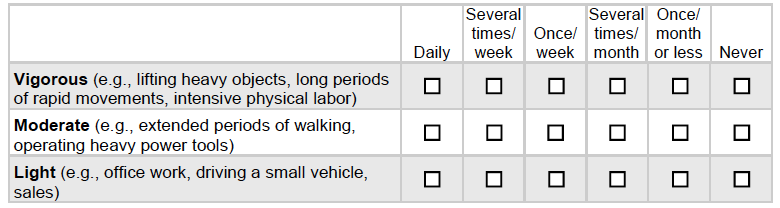

During at home time: *while performing chores in and around your home, how often do you engage in the following levels of activity?*

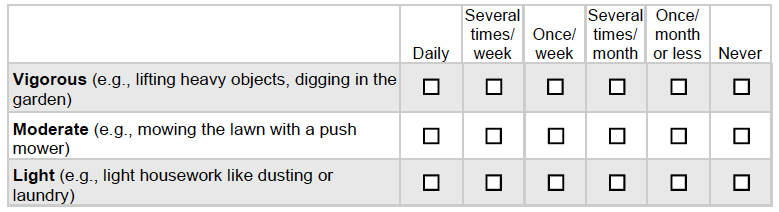

During leisure time: *during your leisure or free time, how often do you engage in the following levels of activity?*

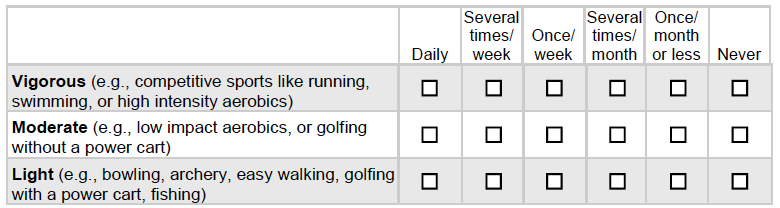

For the All-PA phenotype, the three different levels of PA were summed considering their frequency and weighting intensity to create a total score using the following table

|  | Daily | Several/week | Once/week | Several/month | Once/month | Never |
| --- | --- | --- | --- | --- | --- | --- |
| Vigorous | 15 | 12 | 9 | 6 | 3 | 0 |
| Moderate | 10 | 8 | 6 | 4 | 2 | 0 |
| Light | 5 | 4 | 3 | 2 | 1 | 0 |

After MVP genotyping, which was performed using a customized Affymetrix Axiom Biobank Array and quality control,^3^ EAGLE2^4^ was used for phasing chromosomes and Minimac3 was used for imputation,^5^ using the 1000 Genomes Project reference panel, phase 3, version 5.^6^ Populations were defined using principal component analysis.^1^

For All-PA during leisure time (All-PA-Leisure), we ran independent GWAS for each of three ancestries (EUR; African (AFR); and Admixed American (Latino) (AMR)) (Figure S1). For EUR, we also ran GWAS for vigorous PA during leisure time (Vig-PA-Leisure), work time (Vig-PA-Work), and home time (Vig-PA-Home). We did not have enough power to run GWAS for vigorous PA for AFR and AMR ancestries.

In the quality control procedure for GWAS conducted using PLINK 2.0^7^, we removed variants with imputation quality scores<0.6, Hardy-Weinberg equilibrium *p*-value < 5×10^-5^, minor allele frequency<0.01, missing call rates for variants>0.1, and missing call rates for samples>0.1. Data were aligned to the GRCh37 reference genome. Considering EUR ancestry, after filtering we had 6660206 variants for All-PA-Leisure phenotype, and 6548052, 6536056, 6537158 variants for Vig-PA-Leisure, Vig-PA-Home, and Vig-PA-Work, respectively. For AFR and AMR ancestries, we kept 11984943 and 7969366 variants, respectively (All-PA-Leisure phenotype).

To remove related individuals, we used a threshold of 0.0884 for the kinship coefficients calculated by KING,^8^ resulting in removal of individuals with minimum a second-degree relationship. Then, we implemented an algorithm to optimize keeping the maximum number of individuals with the highest score (for All-PA-Leisure phenotype) or with the highest frequency (for the vigorous activity phenotypes). If two individuals had the same score (or frequency), we would have removed the one with the highest number of relationships. For EUR, we finally had 189,812 individuals for All-PA-Leisure phenotype, and 201,050 for Vig-PA-Leisure, 203,430 for Vig-PA-Home, and 171,278 for Vig-PA-Work. For AFR and AMR ancestries, we kept 27,044 and 10,263 individuals, respectively (All-PA-Leisure phenotype). We ran GWAS analysis using a linear regression model implemented in PLINK 2.0, using sex, age, and the first ten principal components as covariates.

For All-PA-Leisure&Home phenotype, and for All-PA-Leisure&Home&Work, we ran the same QC as described in the main methods, using EUR ancestry. After filtering we had 6678873 variants for All-PA-Leisure&Home, and 6761070 variants for All-PA-Leisure&Home&Work. For the sample size, where we excluded individuals without full information of their PA, we finally had 181,317 individuals for All-PA-Leisure&Home, and 146,929 All-PA-Leisure&Home&Work.

**UKB summary statistics**

UKB is a cohort study of ~500,000 adults between ages 40-69 that live in the United Kingdom (UK) and were recruited from 22 centers across the UK. ^9^ All UKB participants included provided written informed consent.^9^ Ethical approval of the UKB study was given by the North West Multicentre Research Ethics Committee, the National Information Governance Board for Health & Social Care, and the Community Health Index Advisory Group.

From the UKB public databases, we downloaded GWAS summary statistics for the SSOE phenotype with the accession code GCST006100, for moderate-to-vigorous PA (MVPA) with the accession code GCST006097, and for vigorous PA (VPA) with the accession code GCST006098.^10^ Self-reported information were reported via a touchscreen questionnaire. For SSOE, participants needed to answer to the following question: “In the last 4 weeks did you spend any time doing the following?”. Then, there were follow-up questions assessing the frequency and typical duration of “strenuous sports” and of “other exercises”. Cases were defined as individuals spending 2–3 days/week or more doing strenuous sports or other exercises for a duration of 15–30 min or greater, and controls as those individuals who did not indicate spending any time in the last 4 weeks doing either SSOE, obtaining 124,842 cases (*n_case_*) and 225,650 controls (*n_control_*).^10^ We defined the population effective size as *n_eff_*=4/((1/*n_case_*)+(1/*n_control_*)).

The downloaded summary statistics had SNPs filtered according to: Hardy-Weinberg equilibrium *p*-value < 10^-6^, high missingness >1.5%, low minor allele frequency < 0.1%.^10^ We also filtered low imputation quality <0.6, yielding approximately 11.7 million available SNPs.

**Meta-analyses**

METAL^11^ was used for EUR ancestry meta-analysis, which included EUR MVP data for the All-PA-Leisure phenotype and UKB summary statistics for the SSOE phenotype,^10^ in an All-PA-Leisure+SSOE meta-analysis. We also run a cross-ancestry All-PA-Leisure+SSOE meta-analysis, adding AFR and AMR summary statistics from MVP data to the EUR All-PA-Leisure+SSOE meta-analysis (Figure S1). We conducted the meta-analyses using the samplesize method, which considers p-value and direction of effect, weighted according to sample size. This method is recommended when one study used the logistic regression model, such as UKB summary statistics for SSOE phenotype, and another used a linear model, as we did for MVP All-PA-Leisure phenotype, because the effect size from the linear mixed model can be incompatible with the effect size from the logistic regression model.^12^

**X-wide association study (XWAS)**

Sex stratified analysis of the X chromosome was conducted in the MVP sample using XWAS 3.0 on hard call genotypes.^13^ We included 174,375 males and 15,130 females for the All-PA-Leisure phenotype and 229,331 males and 17,072 females for Vig-PA-Leisure. Following sex stratified analysis, test statistics from males and females were combined using Stouffer’s method.^14^ Variants with Hardy-Weinberg equilibrium *p*-value < 5×10^-6^ were filtered, 191,114 variants were included for the All-PA-Leisure phenotype and 191,148 for Vig-PA-Leisure. We used age and the first ten principal components as covariates.

**Annotation, gene-based association analyses, gene-set analyses, functional enrichment, and single-cell expression**

Using the tool MAGMA,^15^ implemented in the FUMA web-based platform,^16^ we annotated variants and ran gene-based tests, gene-set analyses and functional enrichment for each of the two meta-analyses (EUR ancestry and cross-ancestry) for the All-PA-Leisure phenotype and for the GWAS of Vig-PA-Leisure. For annotation we used 1000 Genome Phase3 EUR for EUR ancestry analyses and 1000 Genome Phase3 ALL for the cross-ancestry All-PA-Leisure+SSOE meta-analysis. The other parameters for the identification of lead SNPs were maximum *p*-value cutoff < 0.05, r^2^ threshold to define independent significant SNPs >0.6, the second r^2^ threshold to define lead SNPs >0.1, and the maximum distance between LD blocks to merge into a locus <250 kb. The gene-based test is based on a multiple regression approach to detect gene effects considering SNP *p*-values and linkage disequilibrium. Gene-set analyses are performed for curated gene-sets and GO terms obtained from MsigDB, with terms having a Bonferroni corrected *p*-value<0.05 were considered significant.^17^ Gene property analyses were performed for tissue-specific gene expression using GTEx V8 dataset.^18^

FUMA also includes a tool to test cell type specificity analysis,^19^ which was run using the following datasets of human samples: Allen Brain Atlas Cell Type,^20^ DroNc,^21^ GSE104276,^22^ GSE67835 without fetal samples,^23^ GSE81547,^24^ GSE84133,^25^ GSE89232,^26^ GSE101601 (Linnarsson's lab),^27^ GSE76381 (Linnarsson's lab),^28^ and PsychENCODE.^29^

**SNP-*h*^2^ and genetic correlation.** LDSC^30^ was used to calculate SNP-*h^2^* for EUR ancestry and to calculate genetic correlation^31^ between the EUR meta-analysis of the All-PA-Leisure phenotype and other traits, to quantify genetic similarity (see paragraph “Traits studied for genetic correlation with PA”). To determine which traits were significant we used Benjamini-Hochberg procedure (Table S14, and “Benjamini-Hochberg false discovery procedure”). To estimate SNP-*h^2^* for AFR and AMR cohorts, we calculated LD scores with cov-LDSC^32^ from 10000 random independent individuals for the SNP identified in the HapMap Project^33^.

**Traits studied for genetic correlation with PA**

We estimated the genetic correlation of All-PA-Leisure+SSOE with traits related to cardiovascular disease, respiratory system disease, anthropometric traits, metabolic traits, biological age measures, lifestyle traits, cognitive traits, substance use traits, food consumption, and additional disease traits (Table S30). For these traits, we downloaded EUR-ancestry summary statistics.

Cardiovascular diseases included abdominal and thoracic aortic aneurysm,^34^ coronary artery disease,^35^ heart failure (a clinical syndrome defined by fluid congestion and exercise intolerance due to cardiac dysfunction),^36^ and stroke (which includes ischemic stroke and intracerebral hemorrhage).

Respiratory system diseases included asthma and two traits related to COVID-19, one considering infected individuals versus population (COVID-19), and the other comparing hospitalized cases versus population (COVID-19 hospitalization).^37^

Anthropometric traits included height, bone mineral density,^38^ and body mass index (BMI).^39^

Metabolic traits included high-density lipoprotein (HDL) cholesterol, type 1 diabetes,^40^ type 2 diabetes,^41^ and triglycerides.

Biological age measures included Biological Age Acceleration,^42^ Phenotypic Age Acceleration,^42^ and parental survival.^43^ Biological Age Acceleration and Phenotypic Age Acceleration represent the expected age within the population that corresponds to a person’s estimated mortality risk after accounting for biological or chronological age.^42^ Biological Age Acceleration and Phenotypic Age Acceleration were estimated respectively by the residual of PhenoAge or BioAge after subtracting the effect of chronological age using a linear regression model. PhenoAge and BioAge are two validated biological age predictors.^44,45^ Using biomarker data from the National Health and Nutrition Examination Survey III in the United States, PhenoAge was trained for mortality as a surrogate of biological age, and BioAge was trained for the biological age surrogate of chronological age. PhenoAge is a function of chronological age, albumin, creatinine, C-reactive protein, alkaline phosphatase, glucose, lymphocyte percentage, mean corpuscular volume, red blood cell distribution width, and white blood cell count. BioAge is a function of chronological age, albumin, creatinine, C-reactive protein, alkaline phosphatase, glycated hemoglobin, systolic blood pressure, and total cholesterol.

Lifestyle traits included liking of PA,^46^ household income, wellbeing,^47^ neuroticism, and leisure screen time.^48^

Cognitive traits included educational attainment,^49^ executive functioning,^50^ Alzheimer disease,^51^ and Parkinson’s disease.^52^

Substance use traits included cigarettes per day^53^ and drinks per week.^53^

Food consumption included the behaviors of eating fruit, olive oil, vegetables, and cheese.

Additional disease traits included inflammatory bowel disease,^54^ gastroesophageal reflux disease,^55^ lung cancer,^56^ osteoarthritis, breast cancer,^57^ colorectal cancer,^56^ oral cancer,^56^ and multiple sclerosis.^58^

A report of the study accession is provided in Table S30. Data from the Neale lab were from UKB and downloaded from the website <http://www.nealelab.is/uk-biobank/>.

**Benjamini-Hochberg false discovery procedure**

To determine which traits were significant genetically correlated with PA, we used the Benjamini-Hochberg procedure (Table S14).^59^ We ranked the traits according to their increasing *p*-value, then we calculated the *p*-value’s Benjamini-Hochberg critical value using multiplying each rank by 0.05 and dividing it by the total number of analyzed traits. Finally, the trait *p*-values are compared to the critical values, considering significant the traits until the largest *p*-value was smaller than its corresponding critical value.

**Local genetic correlation**

We ran LAVA^60^ to calculate local genetic correlations between the EUR All-PA-Leisure+SSOE meta-analysis and the 41 traits (Table S30) also studied for genome-wide (global) genetic correlation, to identify shared genetic bases in specific genomic regions. LAVA allows estimation of local genetic heritability and local genetic correlations. We considered 2,495 genomic loci, defined by partitioning the genome into blocks of around 1 Mb and minimizing the LD between the blocks^60^. Based on the number of genomic loci, we defined significant local genetic heritability having a Bonferroni-corrected *p*-value < 0.05/2,495. We evaluated local genetic correlation at loci with pairs of traits already shown to have significant local genetic heritability for both phenotypes. Thus, we conducted 3,088 local genetic correlation tests, defining significant the ones with a Bonferroni-corrected *p*-value < 0.05/3,088.

**Mendelian Randomization**

Mendelian Randomization (MR) analyses allow inferring causality between traits with genetic similarity. TwoSampleMR was performed using four methods: MR Egger, Weighted median, Inverse Variance Weighted (IVW), and Simple Mode.^61^ For the PA trait we used EUR ancestry MVP data of the All-PA-Leisure phenotype, to overpass problems of population stratification and of overlapping samples. We filtered SNPs with *p*-value>10^-5^ and clamping data using the default window of 10000 kb and the r2 cutoff of 0.001. To designate significant results, we applied multiple testing correction for 35 outcomes used both as outcomes and as exposures (*p*-value=7.1×10^-4^). We also used TwoSampleMR to run multivariable MR (MVMR) analyses of All-PA-Leisure phenotype as exposure, to understand the influence that BMI could have as a confounder setting it as exposure. As outcomes, we chose previous significant MR traits which had health importance: abdominal aortic aneurysm, HDL cholesterol, type 2 diabetes, triglycerides, phenotypic age acceleration, gastroesophageal reflux disease, and osteoarthritis. We excluded heart failure from the analyses because of problem of overlapping cohorts. To avoid this problem, for COVID-19 hospitalization we used data from the COVID-19 host genetics initiative without 23andME and MVP data. For the MVMR analyses, we applied to adjust multiple testing (*p*-value=0.0063) because of the 8 outcomes.

**TWAS**

Transcriptome-wide association study (TWAS) are used to identify associated genes to traits. We ran TWAS using FUSION^62^ with the 1000 Genomes LD reference data (https://data.broadinstitute.org/alkesgroup/FUSION/LDREF.tar.bz2) and the GTEx v8 multi-tissue expression weights from 49 tissues (<https://www.mancusolab.com>). These tissues included samples from adipose, adrenal gland, artery, brain, breast, skin, blood, colon, esophagus, heart, kidney, liver, lung, minor salivary gland, muscle, nerve, ovary, pancreas, pituitary, prostate, small intestine, spleen, stomach, testis, thyroid, uterus, and vagina. We performed expression imputation for each autosome using these tissue weights; thus we identified genes that were conditionally independent. Based on these 49 tissue weights and a total of 27,977 genes, we used a multiple testing correction of 3.65×10^-8^. If a gene was significant and expressed in at least two tissues, we chose the gene-expression with the lowest *p*-value.

**Fine-mapping**

FOCUS (Fine-mapping Of CaUsal gene Sets) was used to fine-map TWAS at genomic risk regions, to identify causal genes.^63^ We used the All-PA-Leisure+SSOE EUR meta-analysis and the Vig-PA-Leisure as input GWAS, and the GTEx v7 weights from PrediXcan^64^ combined with Metabolic Syndrome in Men Study (METSIM)^65^, Netherlands Twins Registry (NTR)^66^, Young Finns Study (YFS)^67^, and CommonMind Consortium (CMC)^68^ weights. LD scores were obtained from 1000 Genome phase 3.

We also ran fine-mapping analyses at SNP-level using POLYgenic FUNctionally-informed fine-mapping (PolyFun)^69^. Using precomputed prior causal probabilities based on a meta-analysis of 15 UKB traits allowed extracting per-SNP heritabilities (SNPVAR). These were necessary together with the summary statistics and EUR genotypes from 1000 Genomes Project Phase 3 to perform functionally informed fine-mapping with Sum of Single Effects (SuSiE)^70,71^.

**Multi-Trait Analysis of GWAS (MTAG)**

A valuable method for joint analysis of summary statistics from GWAS of different traits is the multi-trait analysis of GWAS (MTAG)^72^, which can be used to increase power in GWAS analyses. It can be applied to an arbitrary number of traits even if not coming from different samples, generating trait-specific effect estimates for each variant. Here, we ran two MTAG analyses, both with the EUR All-PA-Leisure+SSOE meta-analysis as one trait. First, we joined this trait with leisure screen time.^48^ Because of their negative genetic correlation, we flipped the allele in the LSC statistics. Second, we joined the EUR All-PA-Leisure+SSOE meta-analysis with a measure of PA-liking.^46^

**Enrichment Analysis**

g:Profiler is a toolset which also includes finding biological categories enriched in a gene list.^73^ We performed it on three gene lists resulting from FUMA gene-based test: EUR All-PA-Leisure+SSOE meta-analysis, cross-ancestry All-PA-Leisure+SSOE meta-analysis, and Vig-PA-Leisure.

**Mitochondrial genome association analyses**

We used mitochondria genomes of EUR ancestry individuals from MVP to run mitochondria-SNV association analyses and gene-based association analyses with All-PA-Leisure and Vig-PA-Leisure phenotypes. For the mitochondria-SNV association analyses we filtered missing call rates for variants>0.1, missing call rates for samples>0.1, and excluded monomorphic variants. We thus analyzed 141 SNVs from 189782 individuals for All-PA-Leisure and 142 SNVs from 201018 individuals for Vig-PA-Leisure. For the gene-based association analyses, we assigned the SNV to the mitochondrial genes based on the reference sequence NC_012920.1,^74^ and then we used the R software SKAT v2.2.4 to perform associations with All-PA-Leisure and Vig-PA-Leisure phenotypes, using age, sex, and the 10 first principal components as covariates.^75^ We considered the nominal significant *p*-value (*p*-value≤0.05).

**PheWAS**

We used the Vanderbilt University Medical Center’s (VUMC) EHR database to conduct a phenomewide association analysis (PheWAS) as an empirical exploration of physical activity and its related phenotypes, to investigate a broader range of traits compared to those we used for the genetic correlation analyses.

The VUMC EHR database, also known as the Synthetic Derivative, houses de-identified, longitudinal medical histories of over 3.1 million patients, including their demographic information (age, sex, ethnicity), and clinical information like medications, lab values, procedural and surgical codes and diagnostic codes^76^. Codes from both the International Classification of Diseases 9th and 10th edition (ICD9 and 10) are used by healthcare professionals to code symptoms, procedures and diagnoses in the Synthetic Derivative. A subset of the patients from the Synthetic Derivative also has their biological data associated with their medical records. This subset is referred to as BioVU, and can be used for genomic and phenomic analyses^77-79^.

For our analysis we performed a PheWAS across BioVU records using a polygenic score for physical activity derived from EUR All-PA-Leisure+SSOE meta-analysis. First, we used PRS-CS^80^ to calculate a polygenic score for physical activity for 66,917 BioVU records, of European descent, with genotyped data. Next, using the physical activity polygenic score as the continuous predictor we ran a PheWAS^81^ to explore the phenomic landscape of physical activity. The first step in PheWAS involves mapping related ICD codes to their phecode using the R PheWAS package^82^. The complete list of phecodes and the ICD 9 and 10 codes that are mapped onto them can be found on the PheWAS catalogue (<https://phewascatalog.org/>). Finally, using the EHR driven case-control status for each phecode, we estimated the association between the physical activity polygenic score and the phecodes. We restricted our analysis to 1,254 phecodes that meet three criteria: first, phecodes need over 100 cases or control; second, phecodes should not be specific to a single sex; third, phecodes should not be included in the broader mental health phenotypes. We also controlled for the first 10 ancestry principal components, median age of the record, and reported gender.

**Genomic structural equation modeling**

Genomic structural equation modeling (genomic-SEM) was used to evaluate the overall genetic architecture among the physical activity traits that were included in the EUR GWAS analyses.^83^ Summary statistics for the All-PA-Leisure+SSOE meta-analysis, and for GWAS of Vig-PA-Leisure, Vig-PA-Home, Vig-PA-Work, and liking of PA were used to perform exploratory factor analysis (EFA) and confirmatory factor analysis (CFA). EFA model fit was evaluated by the amount of cumulative variance explained, the strength of sums of square (SS) loadings (SS loadings ≥1), and balance in the proportion of variance explained by each of the individual factors. Traits with factor loadings ≥0.20 in the EFA were allowed to load on the respective factors and evaluated for CFA model fit as determined by conventional fit indices.^83^

**Multi-trait conditional and joint analysis**

Multi-trait conditional and joint analysis (mtCOJO) was performed to study the confounding of socio-economic status in the different kinds of the vigorous physical activities using mtCOJO utility of GCTA.^84^ Socio-economic status was represented with income GWAS summary statistics taken from Neale lab. The conditional analysis was performed individually for each of the three physical activity traits defined in our work: Vig-PA-Leisure, Vig-PA-Home, and Vig-PA-Work. The respective physical activity GWAS summary statistic was used as the target trait and conditioned on the income (covariate trait).

**Survival analysis**

We ran the Cox proportional-hazards model using the package survival 3.3-1 in R, to establish the impact of vigorous PA for the three contexts (leisure, home, work) on risk of dying of any cause. Information was collected in MVP dataset for age, censoring status, and vigorous PA. Age was calculated for censored individuals from the current year minus the year of birth; for dead participants, year of death was provided. The vigorous PA phenotypes present in MVP were previously described in the MVP dataset section of *Supplementary Methods*: we considered Vig-PA-Leisure, Vig-PA-Home, and Vig-PA-Work classification. We had the total information for 252718 individuals (of whom 55849 dead) for Vig-PA-Leisure, 253096 individuals (of whom 55203 dead) for Vig-PA-Home, and 207862 (of whom 43294 dead) for Vig-PA-Work.

### Results

**TWAS and fine-mapping analyses of All-PA-Leisure+SSOE** **meta-analyses**

We performed a TWAS with FUSION to identify gene associations between EUR All-PA-Leisure+SSOE meta-analysis and tissue-specific gene expression data.^62^ We used EUR All-PA-Leisure+SSOE meta-analysis because of the use of EUR linkage disequilibrium (LD) reference data in FUSION. We discovered 80 independent significant genes (Table S16). Running FOCUS fine-mapping analyses following common posterior inclusion probability (PIP) thresholds,^85^ within the 90%-credible set we found 12 with PIP ≥ 0.9 and 19 genes with PIP ≥ 0.7, respectively (Table S17). Fine-mapping analysis at the SNP level (PolyFun) provided 53 SNPs with PIP≥0.95, belonging to 37 genes (Table S18).

**Gene-based association analyses, gene-set analyses, and functional enrichment of cross-ancestry All-PA-Leisure+SSOE meta-analysis**

From gene-based test we obtained 107 significant genes from 19121 protein coding genes (*p*-value=2.6×10^-6^) (Table S9). Like for the EUR meta-analysis, *CADM2* was the strongest significant gene (*p*-value=1.1×10^-18^). The second most significant gene-based association was still on chromosome 3: *CAMKV* (*p*-value=6.6×10^-15^).

Eight significant gene ontology (GO) terms resulted from the MAGMA gene-set analysis (Table S10), with three shared terms between EUR ancestry and cross-ancestry: structural constituent of presynapse, presynaptic active zone organization, and maintenance of presynaptic active zone structure.

MAGMA Tissue Expression Analysis showed four significant enrichments in the brain tissues, with still the two most significant represented in the cerebellar hemisphere and in the cerebellum (Figure S7, S8).

**Mitochondrial genome association analysis of EUR** All-PA**-Leisure**

Mitochondria-SNV association analysis resulted only in a nominally significant marker (rs199951903; *p*-value=0.024; MAF=0.0034) mapping to gene *CYTB*, which encodes Cytochrome b, a component of the mitochondrial electron transport chain. Gene-based association analysis on the mitochondrial genome did not identify any significant results.

**Genetic correlation of EUR All-PA-Leisure+SSOE** **with traits of interest**

Negative genetic correlation was observed with the disease traits, most of anthropometric and metabolic traits (including body mass index (BMI)), the two biological age measures, neuroticism, leisure screen time, and also cigarettes per day. Positive genetic correlations with All-PA-Leisure+SSOE were founded for liking of PA, household income, educational attainment, HDL cholesterol, wellbeing, height, drinks per week, executive functioning, and the analyzed consumed foods: fruit, olive oil, vegetables, and cheese. Genetic correlations were also calculated with other cancer types (breast, colorectal, and oral cancer), Parkinson’s disease, and multiple sclerosis, but none were significant (Table S14).

**MTAG analyses of EUR All-PA-Leisure+SSOE**

We ran an MTAG analysis between the EUR meta-analysis for All-PA-Leisure+SSOE phenotype and leisure screen time, for which we used the EUR ancestry meta-analysis previously reported, which included a total of 526,725 individuals from self-reported data.^48^ For the All-PA-Leisure+SSOE trait, our MTAG analysis provided 74 lead SNPs and 64 genomic risk loci (Table S19). 36 of these lead SNPs were previously not found significant associated to PA, and 59 were previously not found to be significantly associated to leisure screen time. However, only 14 of the 67 previously-identified lead SNPs in the EUR meta-analysis were also significant variants in the MTAG analysis.

We also ran MTAG between the EUR meta-analysis for All-PA-Leisure+SSOE phenotype and liking of PA, given their highest positive genetic correlation (r_g_=0.77±0.02; *p*-value=1.3×10^-210^, *Z*_score_=30.97). Liking of PA is an overall measure derived from 5 PA-liking items in UKB including 151,347 individuals: going to the gym, working up a sweat, exercising with others, exercising alone, and bicycling.^46^ For the PA trait, this analysis provided 59 lead SNPs and 50 genomic risk loci (Table S20). 21 of these lead SNPs were not found previously associated to PA, but only 13 of the previously 67 lead SNPs in the EUR meta-analysis were also maintained significant variants in this MTAG analysis.

The two MTAG analyses, All-PA-Leisure+SSOE with leisure screen time and All-PA-Leisure+SSOE with liking of PA, shared 29 lead SNPs.

**Kinds of vigorous PA and their genetic interrelationships**

Vig-PA-Leisure GWAS identified 8 lead SNPs and 7 genomic risk loci (Figure S11, Table S22). Vig-PA-Work GWAS identified one significant intergenic variant (rs12968236 on chromosome 18) (Figure S12). Vig-PA-Home GWAS did not identify any significant variants (Figure S13).

Thus, we considered Vig-PA-Leisure (SNP-*h*^2^ *Z*_score_=16.34), which had apparent higher power compared to Vig-PA-Work (SNP-*h*^2^ *Z*_score_=5.83) and Vig-PA-Home (SNP-*h*^2^ *Z*_score_=13.65). The highest peak of the Manhattan plot was the variant *SPATS2**rs191602006 on chromosome 12 (*p*-value=1.4×10^-10^), which was also significant in the both previously seen All-PA-Leisure+SSOE meta-analyses of EUR ancestry and cross-ancestry. A sex stratified analysis of the X chromosome conducted on the Vig-PA-Leisure phenotype did not show any significant associated variant (Figure S14).

The gene-based test computed by MAGMA identified 19 significant genes from 18,713 protein coding genes (GWS defined as *p*-value=2.672×10^-6^) (Table S23). As in the All-PA-Leisure+SSOE meta-analyses, *CADM2* was the strongest significant gene (*p*-value=2.0×10^-21^). We did not obtain any significant GO term from MAGMA gene-set analysis. MAGMA Tissue Expression Analysis showed two significant enrichments in the brain tissues: cerebellum and cerebellar hemisphere (Figure S15). FUMA cell type enrichment analysis did not identify significant human cell types across the datasets (see subsection *Single-cell expression* in *Supplementary* *Methods*).

The TWAS performed to link the Vig-PA-Leisure phenotype GWAS to tissue-specific gene expression data identified 5 independent significant genes (Table S24). From the fine-mapping of causal gene sets results, within the 90%-credible set we found one gene (*SLC39A9*) having PIP≥0.9 and two more genes (*IQCH*, *BDH2*) with PIP≥0.7 (Table S25). Fine-mapping analysis at the SNP-level provided 2 SNPs with PIP≥0.95 (Table S26). One of them, *SLC39A8**rs13107325, was also a lead SNP and a genomic risk locus in the MTAG analysis between EUR All-PA-Leisure+SSOE meta-analysis and liking of PA (Table S20).

Functional enrichment analysis using the Vig-PA-Leisure phenotype resulted in a significant term from the protein database CORUM:^86^ the protein complex MAP2K5-PRKCI-SQSTM1 (Table S27).

Mitochondria-SNV association analysis resulted in 5 nominally significant markers (Table S28), with rs199951903 having the lowest *p*-value (*p*-value=0.0084; MAF=0.0034). rs199951903 was also the only nominally significant marker founded in mitochondria-SNV association analysis for All-PA-Leisure (*p*=0.024; MAF=0.0034). Gene-based association analysis found 2 nominal significant associated genes: *TRNV* (*p*-value=0.021, one locus) and *COX3* (*p*-value=0.031, three loci).

**Multi-trait conditional and joint analysis**

We ran conditional analysis for the vigorous PA traits conditioning on income. Compared to previous results without conditioning on income, we only found a significant difference in the heritability of Vig-PA-Leisure (Table S29). Vig-PA-Work and Vig-PA-Home did not show a significant difference in the heritability, as the genetic correlation between each pair of traits (Table S29).

We calculated the genetic correlations between Vig-PA-Leisure, Vig-PA-Work, and Vig-PA-Home individually with the previously analyzed traits of interest, excluding income. Significant statistical differences between Vig-PA-Leisure and Vig-PA-Work were observed for 6 traits, between Vig-PA-Leisure and Vig-PA-Home for 8 traits, and between Vig-PA-Home and Vig-PA-Work for 1 trait (Figure S16).

Supplementary Figures

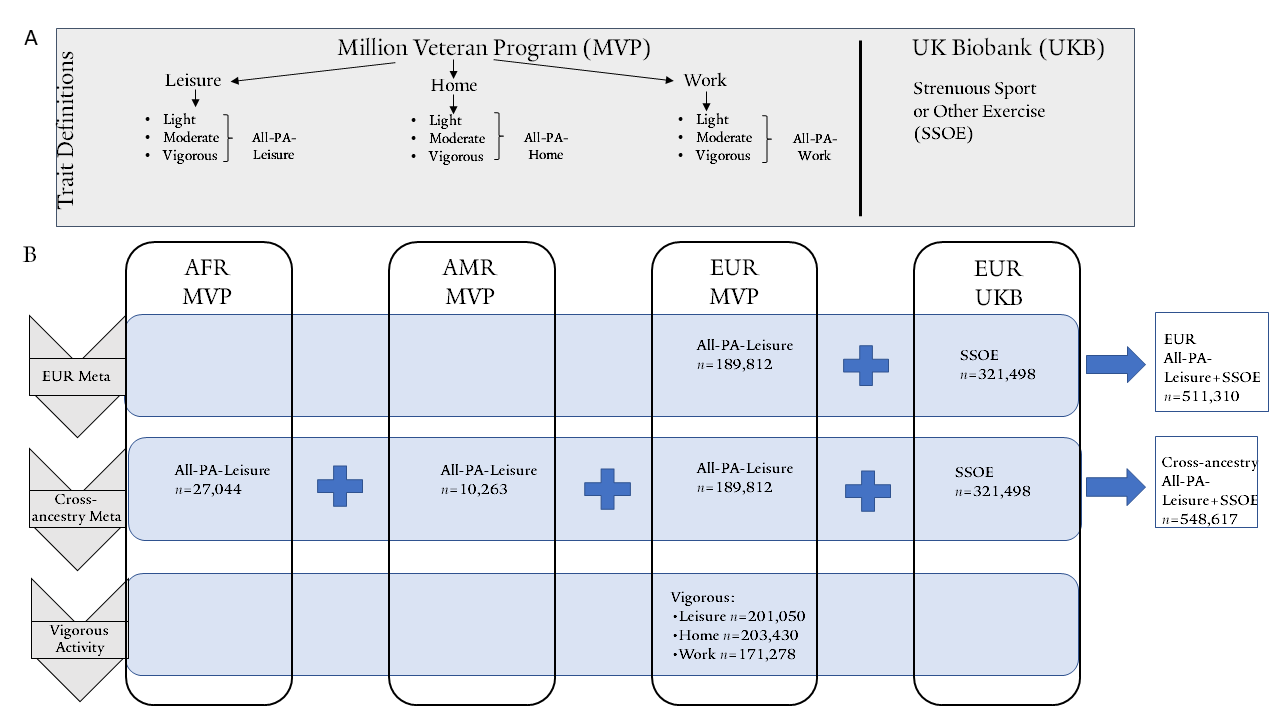

***Figure S1:*** **Overview of the study design.** **Panel A** represents a summary of the physical activity phenotypic information available in the MVP that was used in the current study, along with the UK Biobank data for Strenuous Sport or Other Exercise (SSOE)^10^. **Panel B** depicts the different levels of GWAS performed in the current study, also incorporating information on the respective cohorts and ancestries, and the overall sample size for each trait and resulting meta-analysis.

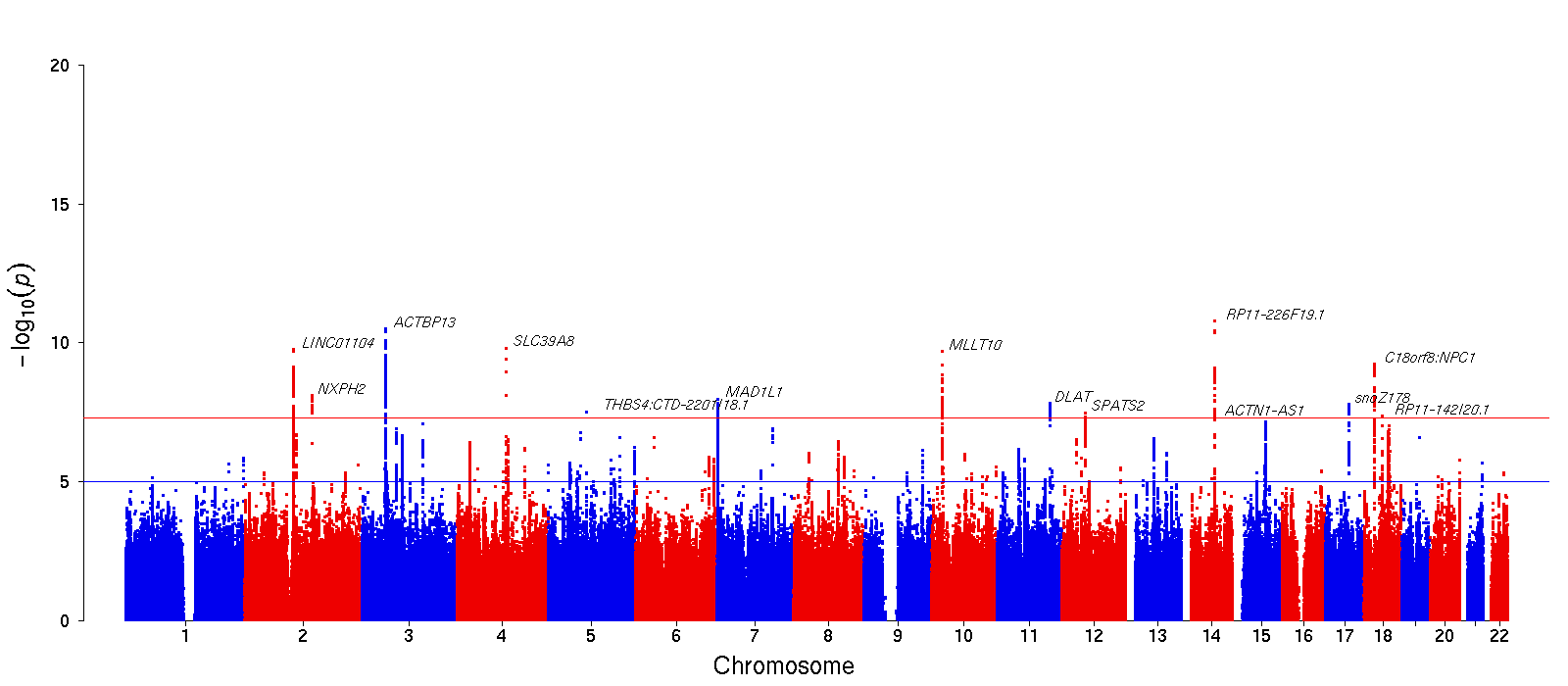

***Figure S2:*** **GWAS analysis of the All-PA-Leisure phenotype in MVP EUR**. Annotated genes are the neareast to each significant lead SNP. The red and blue horizontal lines indicate the genome-wide significance (*p*-value=5×10^-8^) and suggestive significance (*p*-value=10^-5^) levels, respectively.

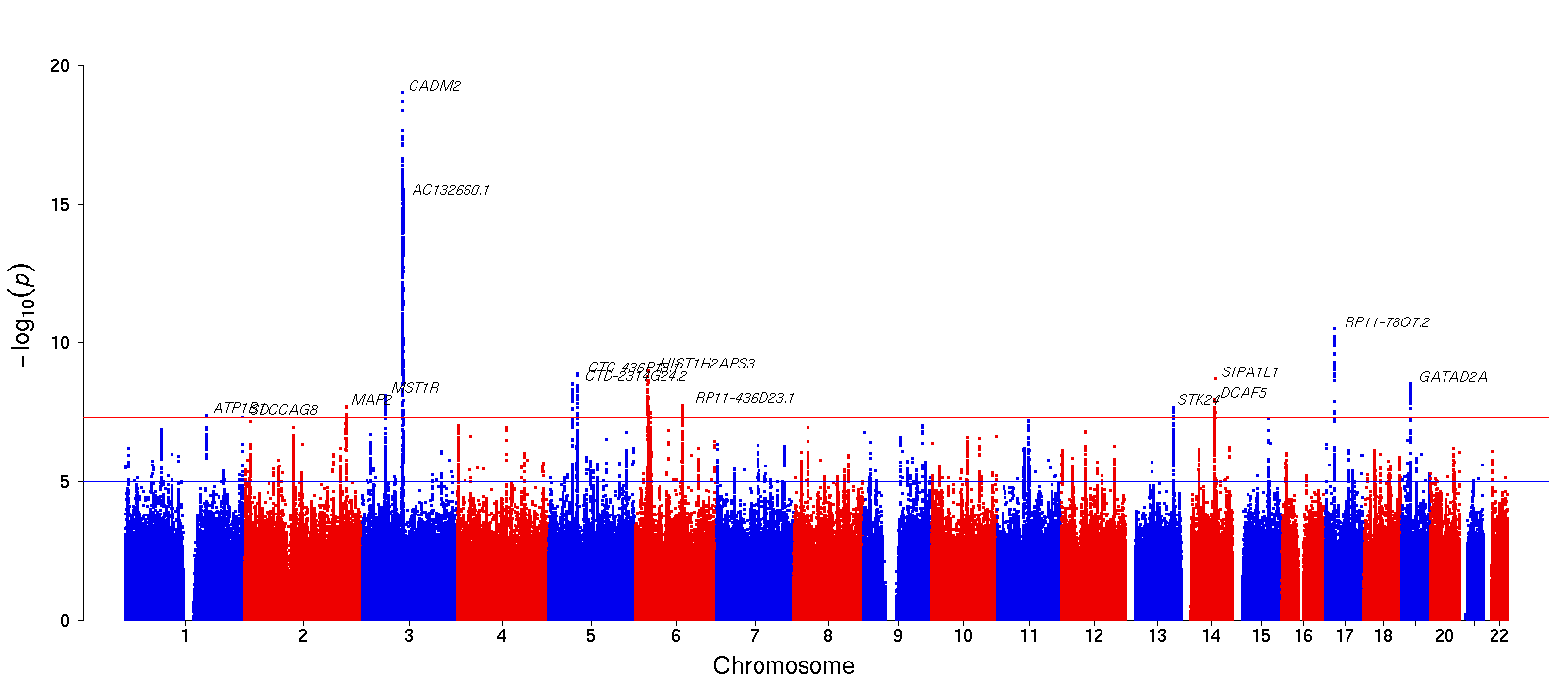

***Figure S3:*** **GWAS analysis of SSOE in UKB.** Annotated genes are the neareast to each significant lead SNP. The red and blue horizontal lines indicate the genome-wide significance (*p*-value=5×10^-8^) and suggestive significance (*p*-value=10^-5^) levels, respectively.

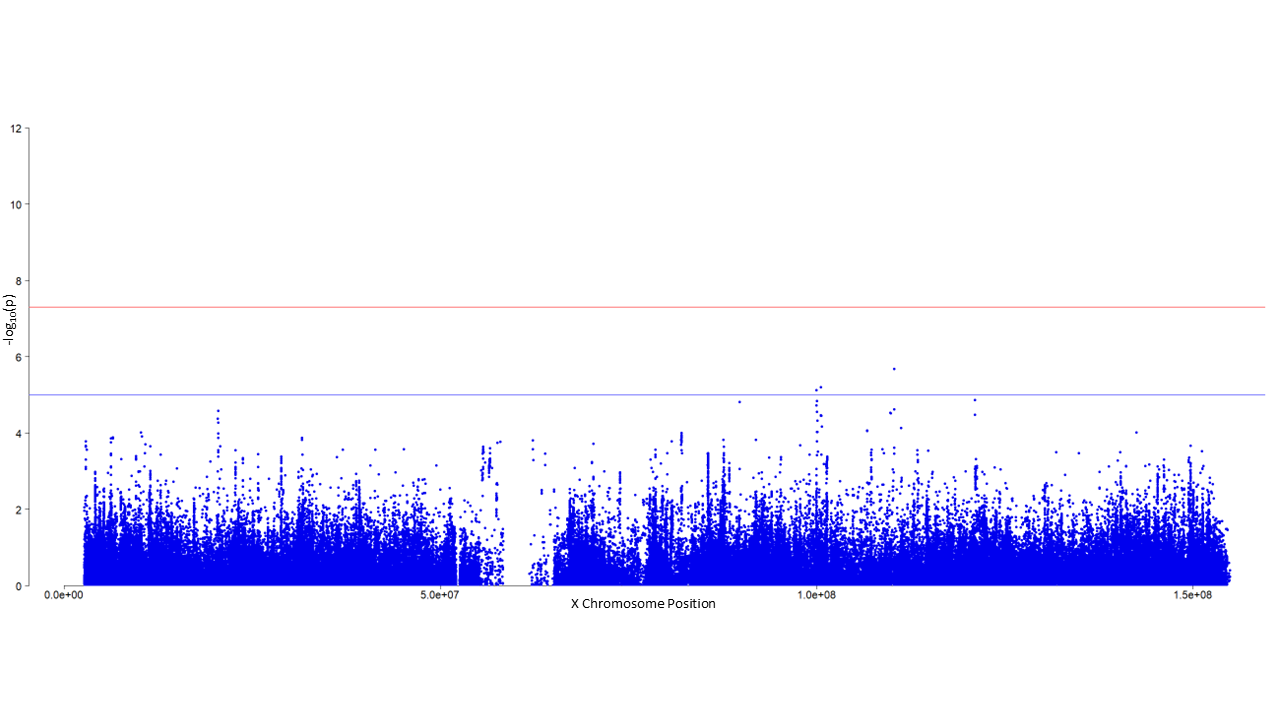

***Figure S4:*** **XWAS of the All-PA-Leisure phenotype for EUR MVP data.** The red and blue horizontal lines indicate the genome-wide significance (*p*-value=5×10^-8^) and suggestive significance (*p*-value=10^-5^) levels, respectively.

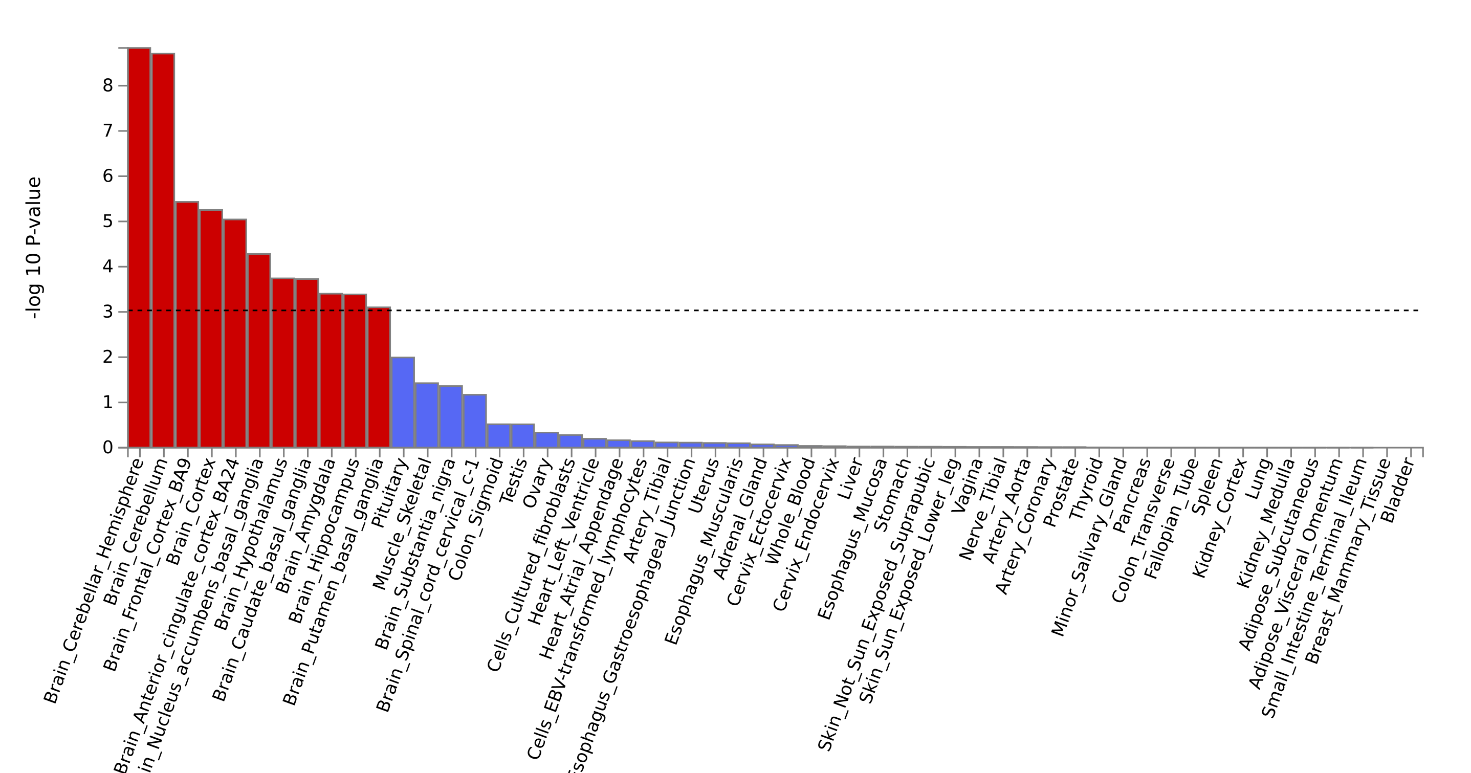

***Figure S5:*** **MAGMA Tissue Expression Analysis with specific tissue** **types** **for EUR All-PA-Leisure+SSOE meta-analysis.**

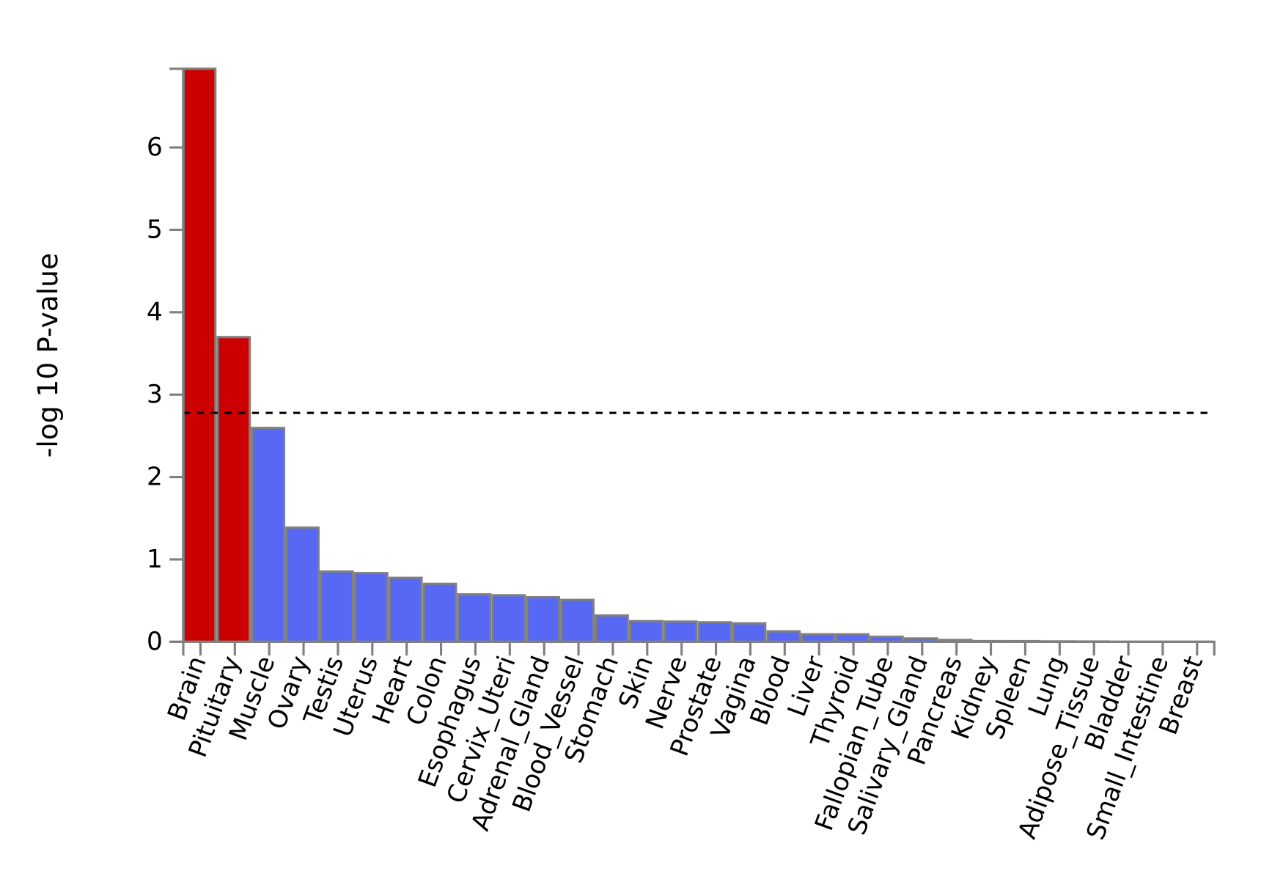

***Figure S6:*** **MAGMA Tissue Expression Analysis with general tissue** **types** **for EUR All-PA-Leisure+SSOE meta-analysis.**

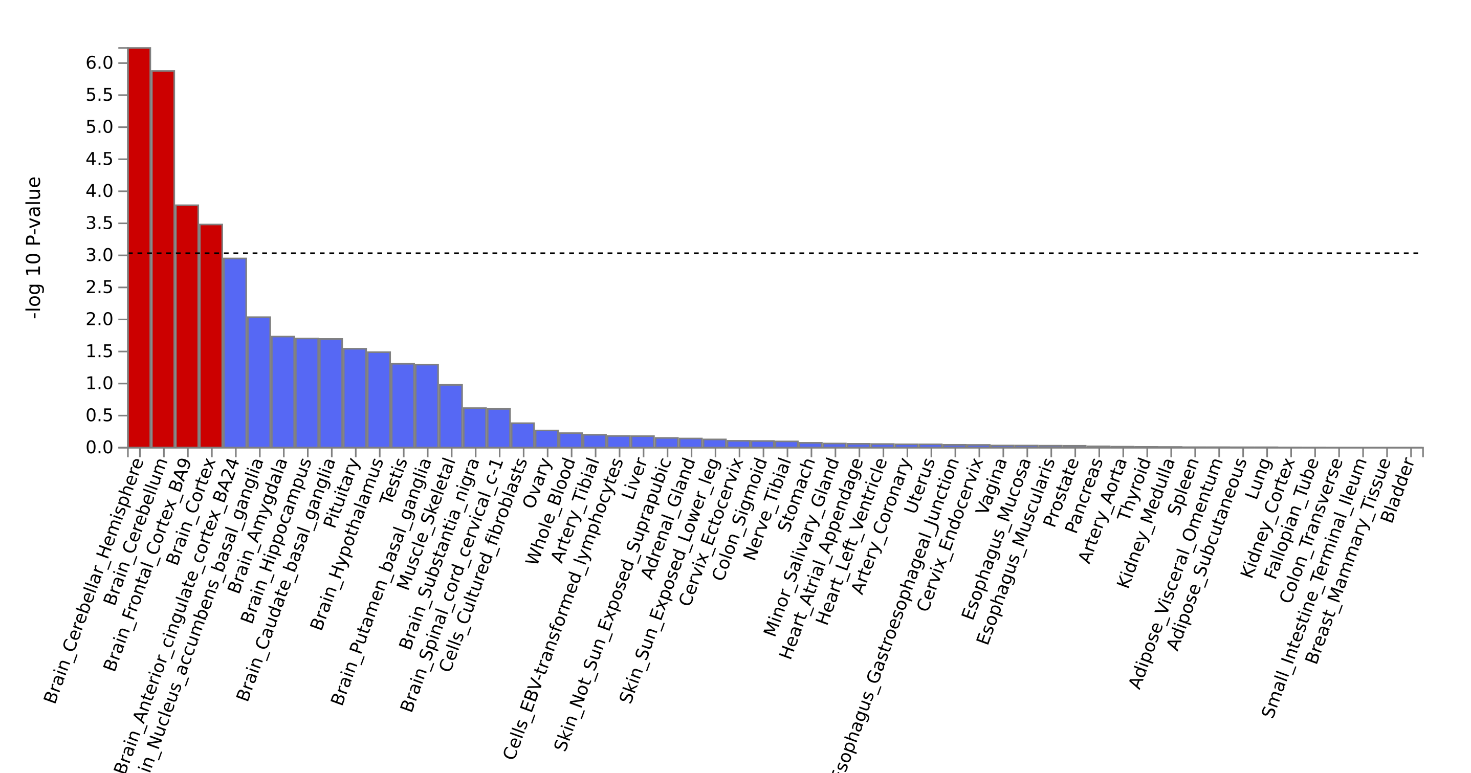

***Figure S7:*** **MAGMA Tissue Expression Analysis with specific tissue types for cross-ancestry All-PA-Leisure+SSOE meta-analysis.**

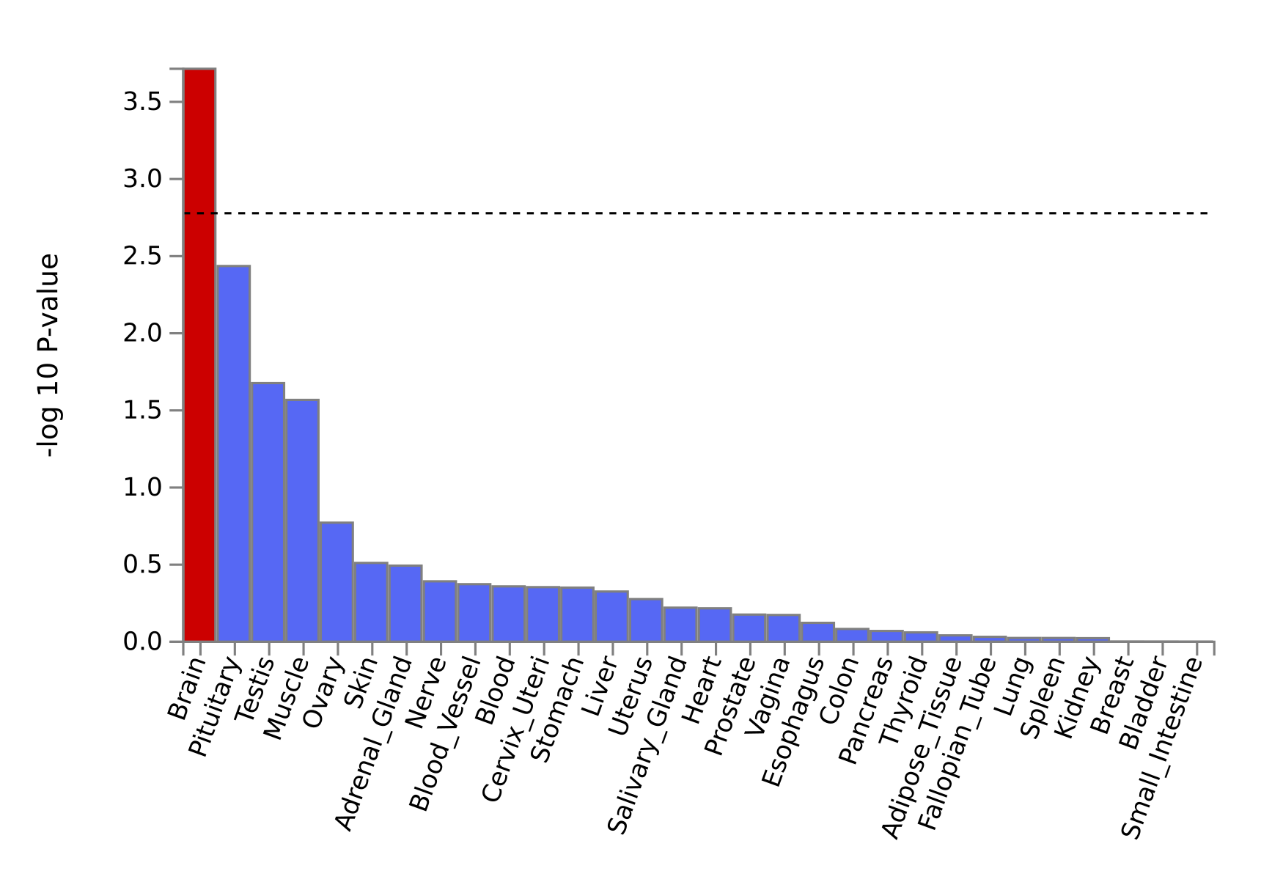

***Figure S8:*** **MAGMA Tissue Expression Analysis with general tissue types for cross-ancestry All-PA-Leisure+SSOE meta-analysis.**

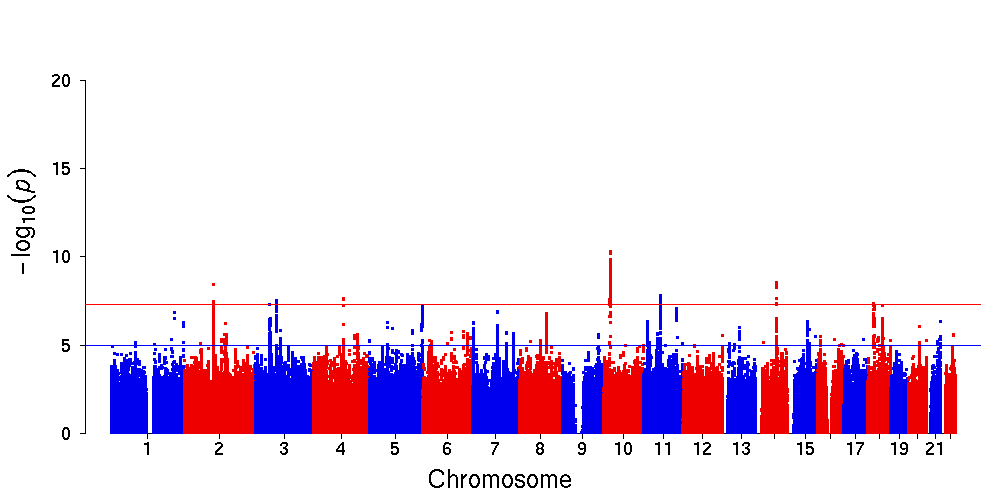

***Figure S9:*** **EUR MVP GWAS analysis of the All-PA-Leisure&Home phenotype.** The red and blue horizontal lines indicate the genome-wide significance (*p*-value=5×10^-8^) and suggestive significance (*p*-value=10^-5^) levels, respectively. We obtained 9 independent significant SNPs, 7 lead SNPs and 7 genomic risk loci.

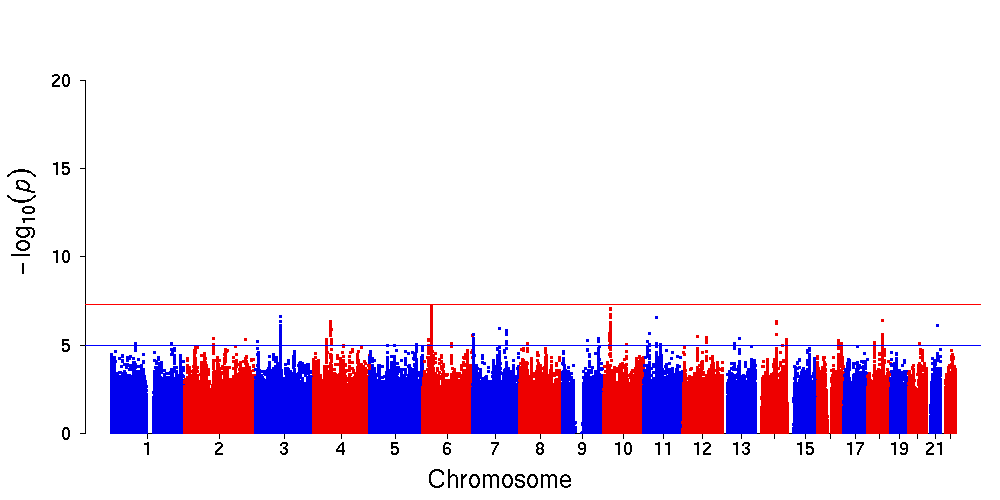

***Figure S10:*** **EUR MVP GWAS analysis of the All-PA-Leisure&Home&Work phenotype.** The red and blue horizontal lines indicate the genome-wide significance (*p*-value=5×10^-8^) and suggestive significance (*p*-value=10^-5^) levels, respectively. No significant variants were found.

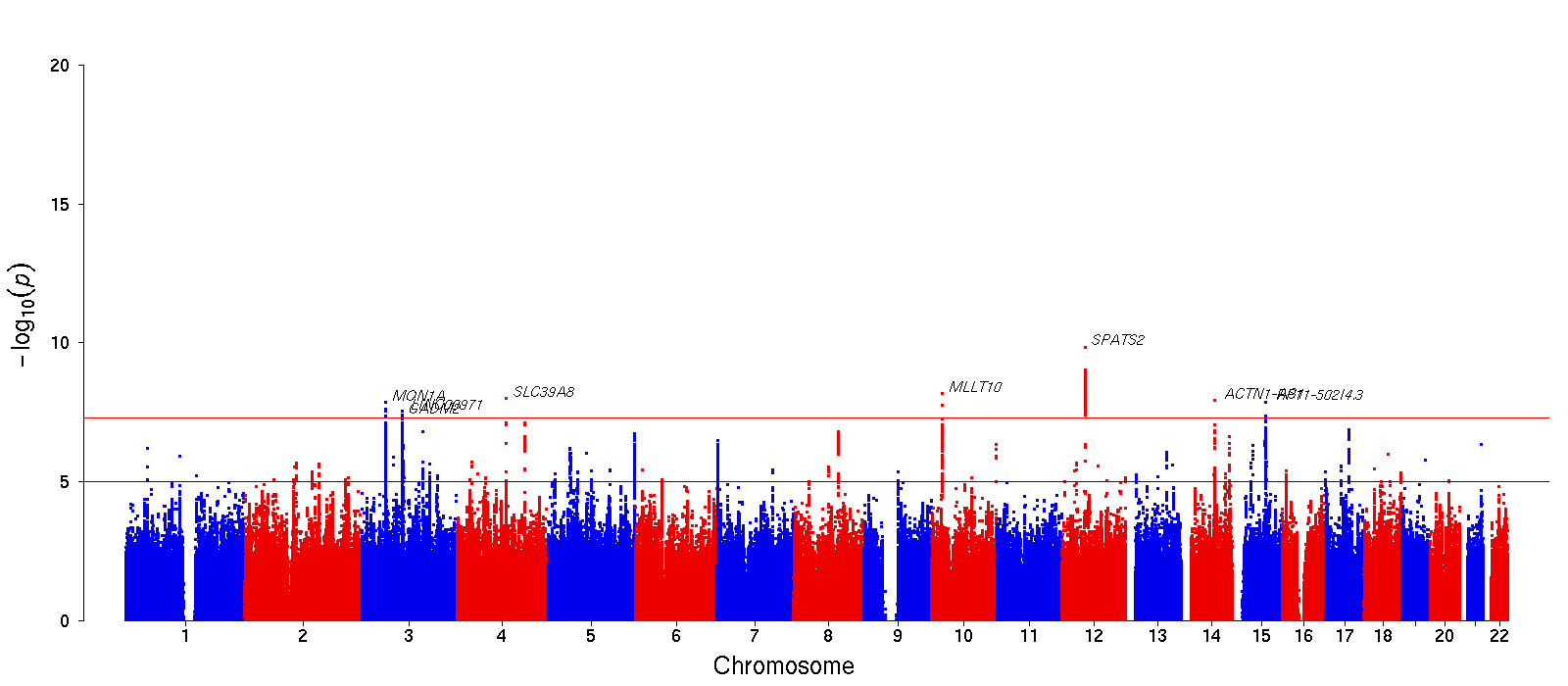

***Figure S11:*** **Manhattan plot of Vig-PA-Leisure.** Annotated genes are the closest to each significant lead SNP. The red and blue horizontal lines indicate the genome-wide significance (*p*-value=5×10^-8^) and suggestive significance (*p*-value=10^-5^) levels, respectively.

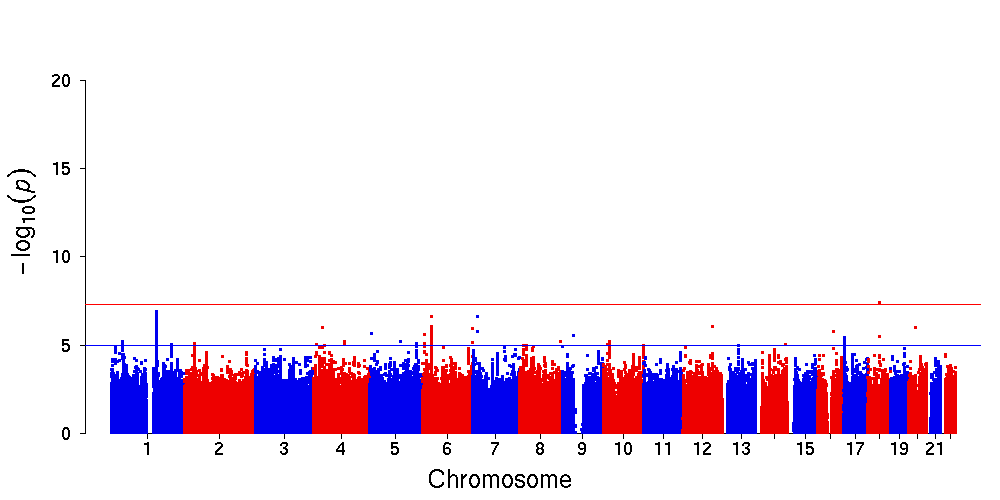

***Figure S12:*** **Manhattan plot of Vig-PA-Work time.** The red and blue horizontal lines indicate the genome-wide significance (*p*-value=5×10^-8^) and suggestive significance (*p*-value=10^-5^) levels, respectively.

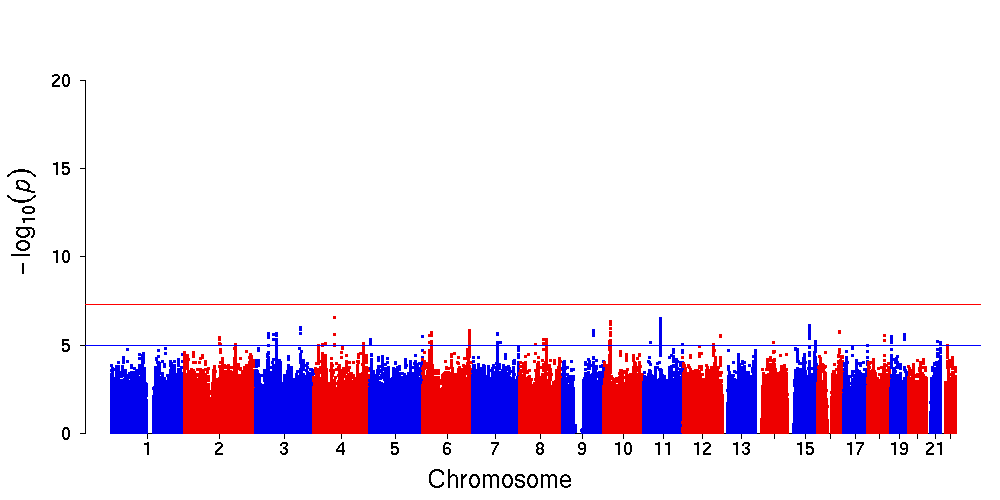

***Figure S13:*** **Manhattan plot of Vig-PA-Home time GWAS.** The red and blue horizontal lines indicate the genome-wide significance (*p*-value=5×10^-8^) and suggestive significance (*p*-value=10^-5^) levels, respectively.

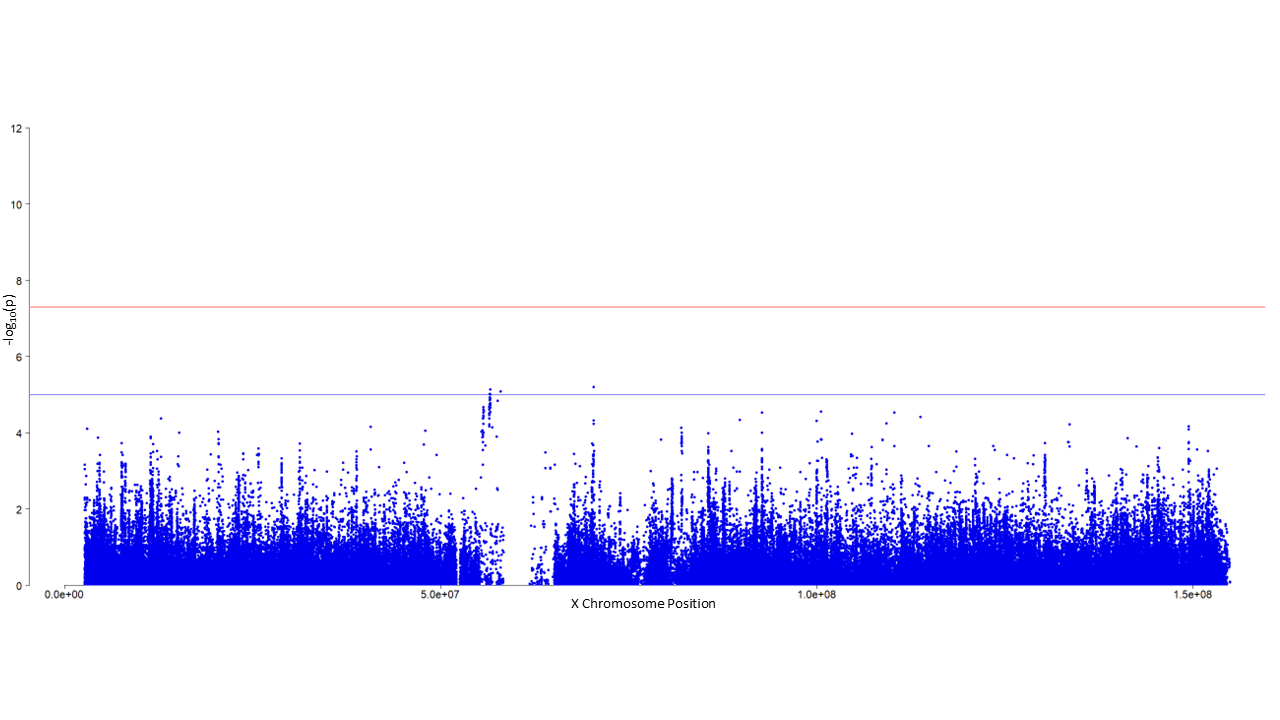

***Figure S14:*** **XWAS of Vig-PA-Leisure phenotype of EUR MVP data.** The red and blue horizontal lines indicate the genome-wide significance (*p*-value=5×10^-8^) and suggestive significance (*p*-value=10^-5^) levels, respectively.

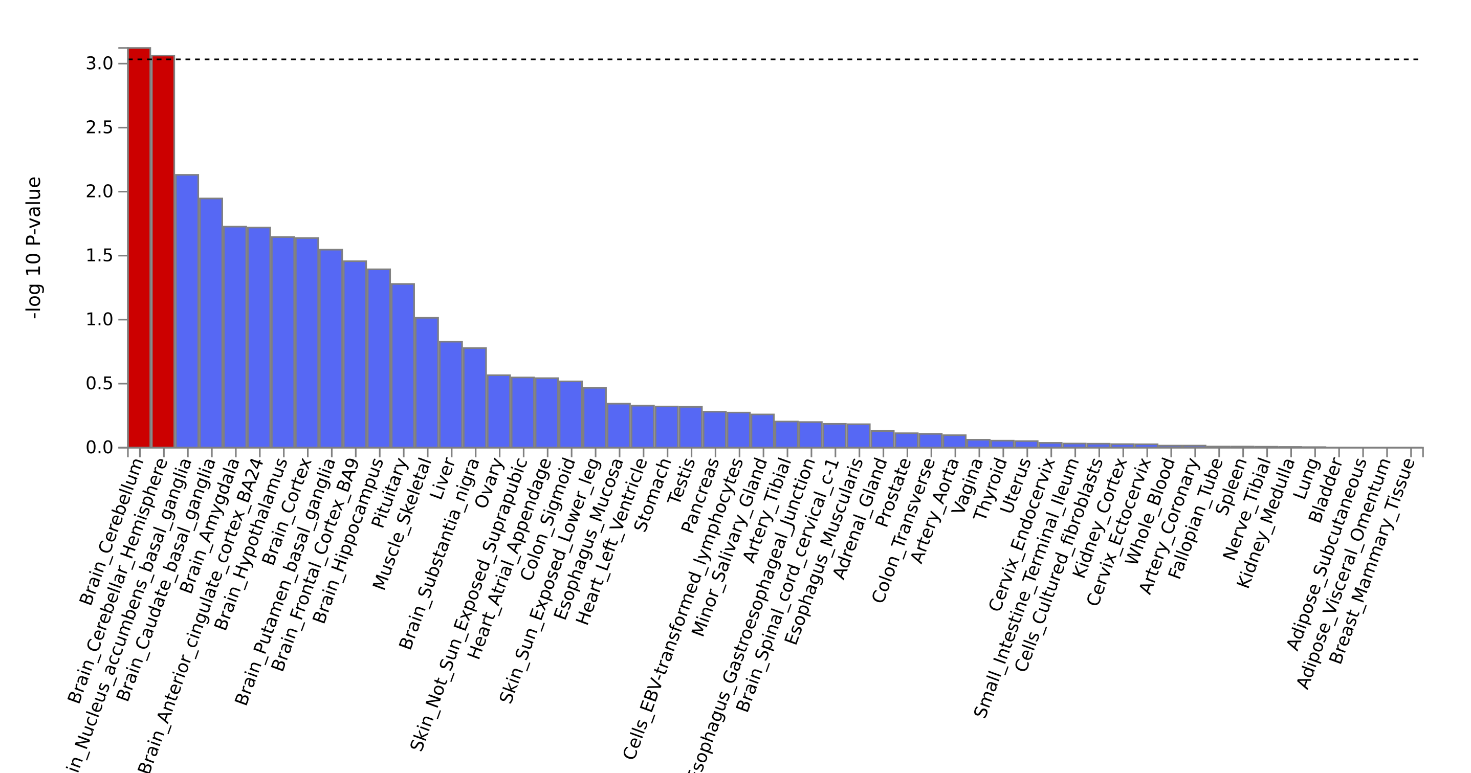

***Figure S15:*** **MAGMA Tissue Expression Analysis with specific tissue types for Vig-PA-Leisure GWAS.**

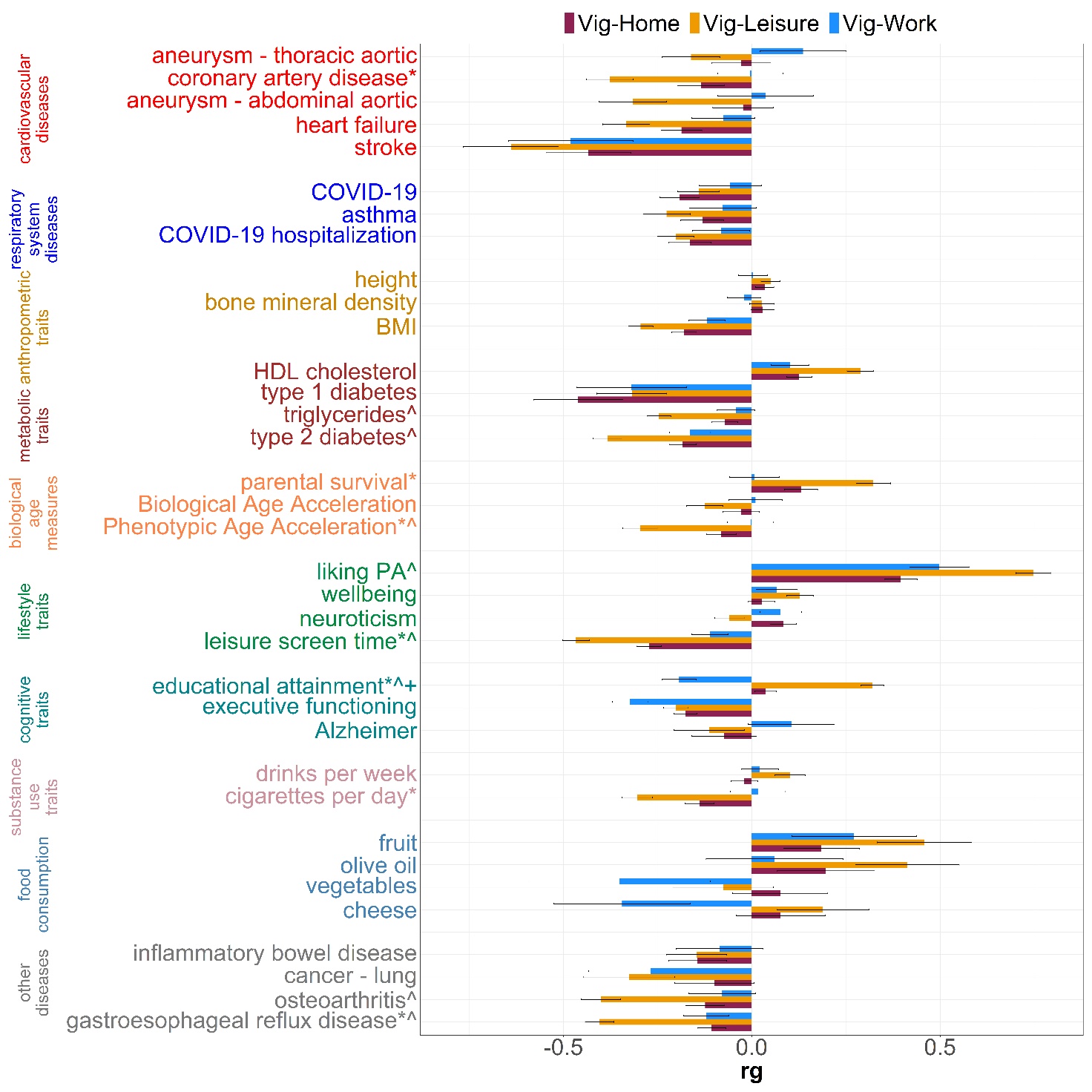

***Figure S16:*** **Genetic correlation between each pair of vigorous PA (Vig-PA-Leisure, Vig-PA-Work, Vig-PA-Home) after conditioning on income, and other traits of interest.** Significantly different correlations were defined having *p*-value < 4.76×10^-4^ after multiple testing correction: * represents significant difference between Vig-PA-Leisure and Vig-PA-Work, ^ between Vig-PA-Leisure and Vig-PA-Home, + between Vig-PA-Home and Vig-PA-Work.
